# Extracellular vesicle miRNA signatures in pediatric onset-multiple sclerosis and obesity-driven immune and metabolic dysregulation

**DOI:** 10.1101/2025.07.16.25331404

**Authors:** Mangesh Dattu Hade, Eduardo Reátegui, J. Nicholas Brenton, Setty M. Magaña

## Abstract

**Background:** Multiple sclerosis (MS) is a chronic, demyelinating autoimmune syndrome of the central nervous system (CNS). The cause of MS remains unknown; however, a dysregulated metabolome appears to play a contributing role. Up to 10% of all MS cases occur in childhood or adolescence. Childhood obesity is an independent risk factor for the development of MS. Pediatric-onset MS (POMS) provides a unique opportunity to study obesity-associated autoimmunity in the absence of age-related comorbidities. Circulating extracellular vesicle-encapsulated microRNAs (EV-miRNAs) have emerged as stable biomarkers and regulators of immune and metabolic pathways in neuroinflammatory disorders but have not been investigated in POMS.

**Methods:** We performed bulk and single EV analysis on platelet-poor plasma EVs from children with POMS and age-, sex-, and body mass index (BMI)-matched controls. EVs were isolated via tangential flow titration (TFF) and subjected to miRNA-Seq. Differential expression analyses were performed to identify EV-miRNAs unique to MS and obesity. Functional enrichment was employed to elucidate impacted biological pathways. Key findings were cross-referenced with known and published pathways. Single EV analysis using total internal reflection fluorescence microscopy (TIRFM) was employed to identify EV and obesity-specific markers at single-particle resolution.

**Results:** Bulk EV characterization confirmed the presence of small EVs with classic biophysical characteristics. Particle size was comparable across both groups and ranged between 70 to 91 nm and concentration range was 2 x 10^10^ to 1 x 10^11^/ mL. EV-miRNA sequencing demonstrated marked global differential transcriptomics between POMS and controls, irrespective of BMI. Notably, we identified MS-enriched EV-miRNAs, such as miR-29b-3p, miR-4326, miR-671-3p, and miR-139-3p, that are involved in cholesterol homeostasis, inflammatory signaling, T-cell regulation and blood-brain-barrier (BBB) integrity. These MS-specific alterations implicate EV-miRNA cargo in modulating pathobiologically relevant pathways, such as immune regulation, neuroinflammation, and lipid metabolism. Obesity also had a pronounced, group-specific impact on EV-miRNAs. Subgroup analysis comparing controls to obese POMS revealed alterations in EV-miRNAs involved in regulating metabolism homeostasis, pro-adipogenesis and inflammation (e.g., miR-29a-3p, miR-142-5p, miR-29c-3p, miR-27a-3p). In POMS patients, obesity further amplified these trends with markedly increased pathogenic EV-miRNAs, including miR-142-5p, miR-29a-3p, and let-7d-5p, and significant suppression of protective EV-miRNAs, such as miR-30d-5p, let-7c-5p, and miR-151a-5p when comparing normal weight MS versus obese MS. Enrichment analysis revealed dysregulated EV-miRNAs in MS target pathways central to immune activation, neuroinflammation, and lipid metabolism, while obesity accentuated disturbances in pathways, for example AMPK signaling, insulin resistance, and axon guidance. Single EV analysis via TIRFM confirmed the presence of obesity-specific EVs within the circulating plasma EV pool of POMS.

**Conclusion:** EV-miRNA profiles differ significantly between POMS patients and controls, reflecting aberrant immune and metabolic regulation in disease. Furthermore, obesity resulted in an expanded MS-related, EV-miRNA dysregulated repertoire, underscoring a prospectively novel cellular mechanism underlying obesity-associated CNS autoimmunity. These findings highlight plasma EVs as promising minimally invasive biomarkers in POMS and provide novel therapeutic candidates for future validation in larger cohorts.

## Introduction

Multiple sclerosis (MS) is a chronic autoimmune disorder of the central nervous system (CNS) characterized by increased blood-brain-barrier (BBB) permeability, immune-mediated demyelination, and axonal injury[1–3]. While MS is typically diagnosed in adulthood, pediatric-onset MS (POMS) accounts for up to 10% of all cases and poses unique challenges due to its early onset and impact on CNS development [4–8]. Despite its clinical relevance, POMS remains underexplored at the molecular level, limiting the discovery of biomarkers and therapeutic targets tailored to this population.

Childhood obesity, an independent risk factor for POMS [9,10], is present years prior to the first clinical manifestation(s) of MS, suggesting that excess adiposity may contribute to CNS autoimmunity [9,11,12]. Mechanistically, obesity induces a chronic low-grade inflammatory state characterized by elevated leptin, IL-6, and TNF-α, along with decreased anti-inflammatory adipokines such as adiponectin [13–15]. This inflammatory milieu perturbs immune tolerance and may exacerbate autoreactive T and B cell responses [13–15].

A growing body of research implicates microRNAs (miRNAs) as critical modulators of immune tolerance, neuroinflammation, and myelination in MS [16–18]. MiRNAs are small noncoding transcripts that post-transcriptionally modify gene expression by binding to complementary sequences on target messenger RNAs (mRNAs), leading to mRNA degradation or translational repression Dysregulated miRNAs, such as miR-29b and the let-7 family, are important modulators of numerous MS pathogenic pathways, including the IFN-γ–T-bet axis, Th17/Treg homeostasis, and blood-brain-barrier integrity [19,20] [16]. Limited studies have investigated the role of circulating miRNAs in POMS [21].

Recent attention has turned to EV-miRNAs as potential biomarkers and mechanistic drivers of MS [22] . EVs, such as exosomes, are lipid bilayer-enclosed nanoparticles, which provide protection against RNA degradation, and are secreted by virtually all cells and found in all biofluids [23]. EVs represent a novel mechanism of intercellular cross-talk by transmitting bioactive cargoes, such as miRNAs, proteins, and lipids, to distant and local recipient cells [24,25]. Because EVs reflect the activation status of the secreting parental cell, EV-derived miRNAs have emerged as noninvasive ‘liquid biopsies’ of CNS processes and functional mediators of neuroimmune pathomechanisms. In adult MS, distinct EV-miRNA profiles have been reported in serum and CSF, correlating with disease subtype and activity [24–26]. For instance, miR-146a and miR-21, encapsulated in EVs, have been shown to distinguish relapsing and progressive MS and reflect immune status during relapse [27]. Moreover, EVs from dendritic cells containing miR-155 or miR-146a can be transferred to recipient immune cells, promoting or attenuating inflammatory gene expression, respectively [24]. These findings highlight the dual role of EV miRNAs as both biomarkers and intercellular signaling molecules in MS.

Importantly, obesity is also associated with changes in EV cargoes, including miRNAs. Studies in obese adults have demonstrated that plasma EVs are enriched in miRNAs that regulate lipid metabolism, insulin sensitivity, and inflammatory pathways [28]. In a mouse model, exosomal transfer of obesity-associated miRNAs induced metabolic syndrome in lean animals, establishing a causal role for EV-miRNAs in disease propagation [28]. These findings raise the possibility that in obese POMS patients, EV-miRNAs may serve as a molecular conduit linking peripheral metabolic dysfunction with CNS autoimmunity.

To date, no studies have characterized EV-miRNA profiles in POMS, nor examined how comorbid obesity may modulate this molecular landscape. This represents a critical gap in understanding the molecular underpinnings linking peripheral adiposity with CNS autoimmunity. In this pilot study, we conducted the first comprehensive profiling of the circulating EV-miRNome in youth with POMS compared to age-, sex-, BMI-matched controls. Our objectives were to investigate an MS-specific EV-miRNome, to delineate the impact of obesity on EV-miRNA signatures, and to posit on pathobiologically relevant pathways through functional enrichment analysis. We found distinct MS-specific EV-miRNA repertoires, independent of obesity, that distinguished subjects with POMS from controls. Additionally, our results implicate unique obesity-driven EV-miRNAs that amplify dysregulated pro-versus anti-inflammatory responses, cholesterol metabolism, and neuroinflammatory signaling [29–31]. Our findings suggest that EV-miRNAs may serve as disease-specific biomarkers of POMS and lay the groundwork for future mechanistic studies into the molecular underpinnings of early-life adiposity and MS pathogenesis.

## Results

### Enrolled study participants

For this pilot study, we utilized banked plasma from eleven subjects (MS, n=5; controls, n=6) that were previously enrolled into the Body Composition in POMS study (NCT04593082) at the University of Virginia (UVA). POMS subjects met diagnostic criteria [32] for relapsing-remitting MS, with onset prior to the age of 18 years. The POMS cohort had to be within 3 years of MS diagnosis for inclusion in this study. Controls were age (+/- 1 year), sex-, and BMI category-matched to POMS subjects and were healthy individuals without a diagnosis of MS or any other autoimmune disease. All subjects had blood samples collected during early morning and while in the fasting state, with at least 8 hours of fasting prior to sample collection. The age range of participants was 16–20 years.The control cohort had an age range of 16 to 20 years and a BMI range of 23 to 48.5 kg/m². The POMS cohort had an age range of 16 to 19 years and a BMI range of 21.6 to 35.4 kg/m² (**Table 1**).

**Table 1:** Participant demographics.

| ID | Sex | Ethnicity | BMI (kg/m<sup>2</sup>) | Group designation |
| --- | --- | --- | --- | --- |
| HC-NW-01 | M | Non-Hispanic | 24.9 | Control, Normal weight |
| HC-NW -02 | M | Non-Hispanic | 23 | Control, Normal weight |
| HC-NW -03 | F | Non-Hispanic | 25 | Control, Normal weight |
| HC-OB-10 | M | Non-Hispanic | 29.4 | Pediatric Overweight/Obese |
| HC-OB-11 | F | Non-Hispanic | 27 | Pediatric Overweight/Obese |
| HC-OB-13 | F | Non-Hispanic | 48.5 | Pediatric Overweight/Obese |
| MS-NW-01 | M | Non-Hispanic | 21.6 | POMS, Normal weight |
| MS-OB-02 | M | Non-Hispanic | 35.4 | POMS, Overweight/Obese |
| MS-OB-05 | F | Non-Hispanic | 32.4 | POMS, Overweight/Obese |
| MS-OB-06 | F | Non-Hispanic | 34.2 | POMS, Overweight/Obese |
| MS-NW-12 | M | Hispanic | 24.3 | POMS, Normal weight |

### Heterogeneous circulating EVs in matched POMS patients and controls

We employed our previously established and validated pipeline for the isolation and comprehensive characterization of circulating EVs[33,34] from POMS patients and matched controls (**Figure 1A**; see Materials and Methods).

**Figure 1.**
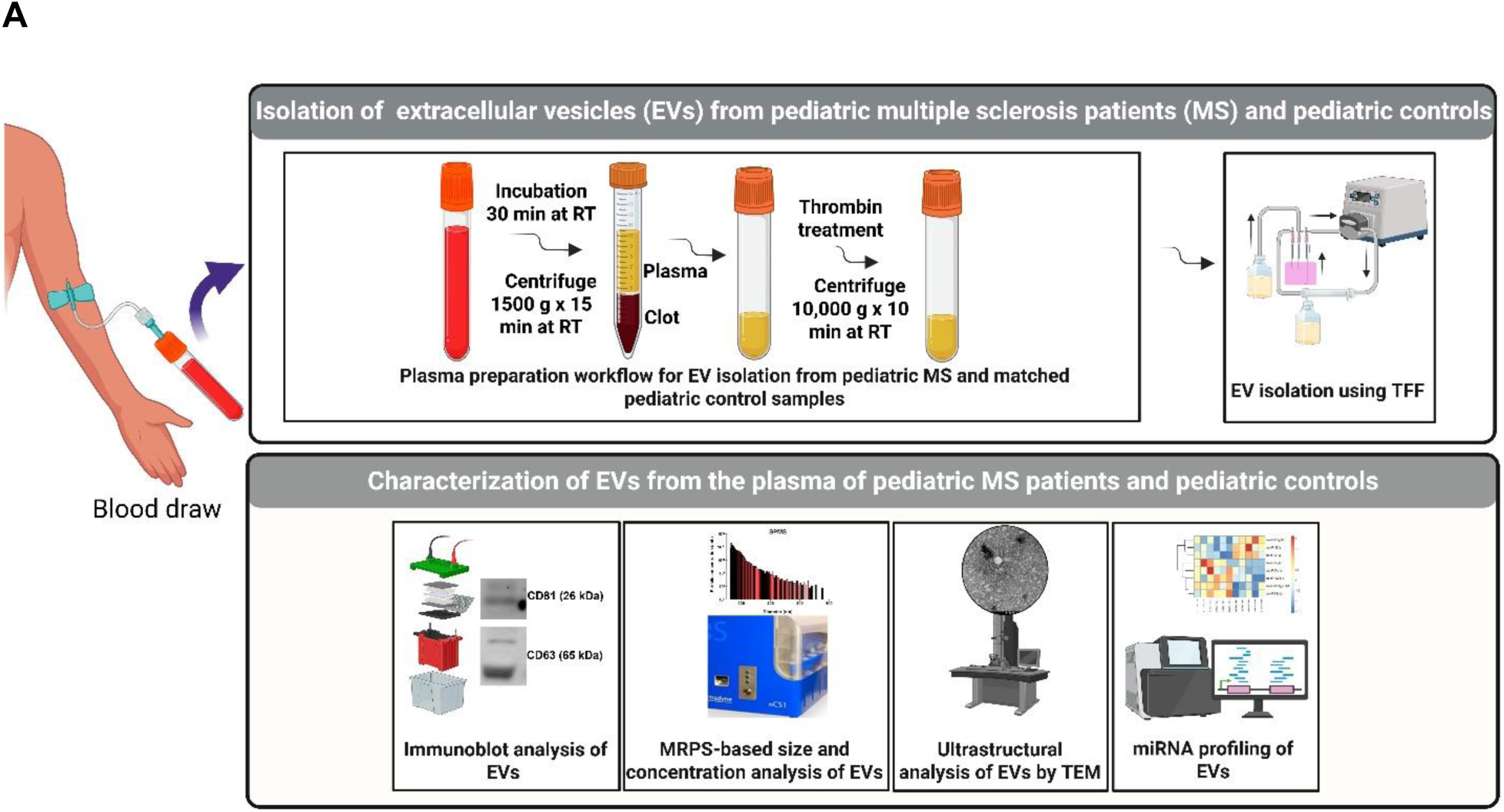
Isolation and multiparametric characterization of circulating EVs from POMS patients and matched controls. (A) EV isolation workflow: Whole blood was collected and incubated at room temperature (RT) for 30 min, followed by centrifugation at 1500 × g for 15 min to separate plasma. Plasma was defibrinated using thrombin and centrifuged again at 10,000 × g for 10 min at RT to remove residual fibrin. EVs were subsequently isolated from defibrinated plasma via tangential flow filtration (TFF). Characterization steps included immunoblot analysis for EV markers (CD81, CD63) and the non-EV marker CANX, particle sizing and concentration analysis using microfluidic resistive pulse sensing (MRPS), ultrastructural assessment via transmission electron microscopy (TEM), and miRNA profiling. **(B) Detailed characterization of circulating EVs from POMS patients and matched controls**. (a) MRPS analysis demonstrates comparable EV particle size distributions between POMS and controls, with median particle diameters of approximately 81.8 nm (POMS) and 71.7 nm (controls). Data represent three independent biological replicates per group. (b) Representative TEM images illustrating heterogeneous EV populations exhibiting characteristic cup-shaped morphology consistently across both groups. (n=3 independent biological replicates) Scale bar = 100 nm. (c) Immunoblot analysis confirmed heterogeneous expression of classical EV markers CD81 (26 kDa), CD63 (65 kDa) and the absence of the non-EV marker Calnexin (97 kDa) in EV samples from POMS and controls plasma (n=3 independent biological replicates). MRPS, microfluidic resistive pulse sensing; TEM, transmission electron microscopy.

To validate EV integrity and quality, isolated EVs were rigorously characterized using multiparametric approaches including microfluidic resistive pulse sensing (MRPS), transmission electron microscopy (TEM), and immunoblotting. MRPS analysis revealed similar and consistent particle size distributions in EVs from both POMS patients and control groups, with median particle diameters of approximately 81.8 nm for POMS and 71.7 nm for controls (**Figure 1B**).

Ultrastructural examination via TEM confirmed the typical ‘cup-shaped’ morphology of EVs, revealing heterogeneous polydispersed populations of small vesicles (**Figure 1Bb**). Additionally, immunoblot analysis confirmed expression of canonical EV markers CD81 (26 kDa) and CD63 (65 kDa), and the absence of the non-EV marker Calnexin (97 kDa). (**Figure 1Bc**), confirming our particles as EVs and not co-isolates.

Collectively, these results highlight the successful isolation and biophysical characterization of plasma-derived EVs from POMS and control subjects.

### MS-specific EV-miRNA signature distinguishes POMS from matched controls

To investigate disease-specific EV-miRNA repertoires, we performed differential expression analysis. A total of 799 miRNAs were identified, and miR-4326_R+3 and miR-29b-3p_L+1R-1 miRNAs were significantly dysregulated in MS plasma EVs compared to controls. Given the exploratory nature of this study and the small sample size, we applied a less stringent statistical threshold (*p* < 0.1) to define significant miRNAs (Supplementary Table 1), while also evaluating miRNAs at more conventional thresholds of *p* < 0.001, *p* < 0.05 and *p* < 0.01 (Table 2). A total of 465 miRNAs were detected across MS and control groups. Among them, 399 miRNAs were shared between both groups, while 36 miRNAs were unique to MS patients and 30 were exclusive to controls (**Figure 1A**). Volcano plot analysis (**Figure 1B**) provides a global view of miRNAs upregulated (red) and downregulated (blue) in MS versus controls. Labeled miRNAs are significantly differentially expressed between MS and controls. Hierarchical clustering of differentially expressed miRNAs revealed distinct heatmap expression signatures amongst the two groups, suggesting MS-specific miRNA candidates (**Figure 1C**). Notable dysregulated miRNAs are listed in **Table 2**. Correlation analysis demonstrated similar expression for most miRNAs in both groups, but labeled miRNAs (e.g., miR-26a-5p, miR-142-5p, and miR-128-3p) were significantly elevated and may point to pathobiologically relevant miRNAs (**Figure 1D**).

**Table 2.** Top differentially expressed EV-miRNAs in Controls vs. POMS.

| Downregulated DEMs | FC | log2(FC) | *p-value* * |
| --- | --- | --- | --- |
| miR-671-3p | 0.63 | -0.67 | 7.67E-03 |
| miR-139-3p | 0.47 | -1.10 | 1.30E-02 |
| miR-93-3p_R+1 | 0.70 | -0.51 | 1.33E-02 |
| Upregulated DEMs |  |  |  |
| miR-4326_R+3 | 2.80 | 1.49 | 2.03E-02 |
| miR-29b-3p_L+1R-1 | 1.59 | 0.67 | 8.51E-03 |

DEM, Differentially expressed miRNAs; FC, fold change; POMS, pediatric-onset multiple sclerosis

**Figure 1.**
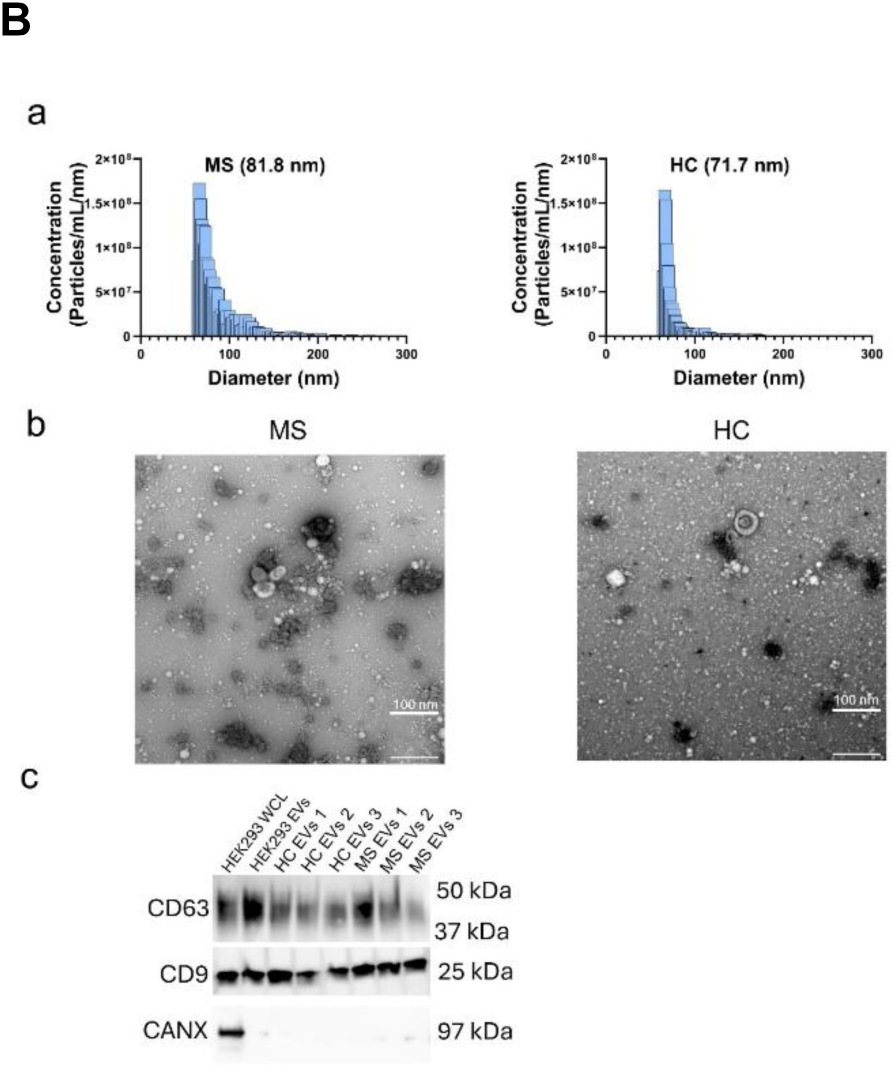
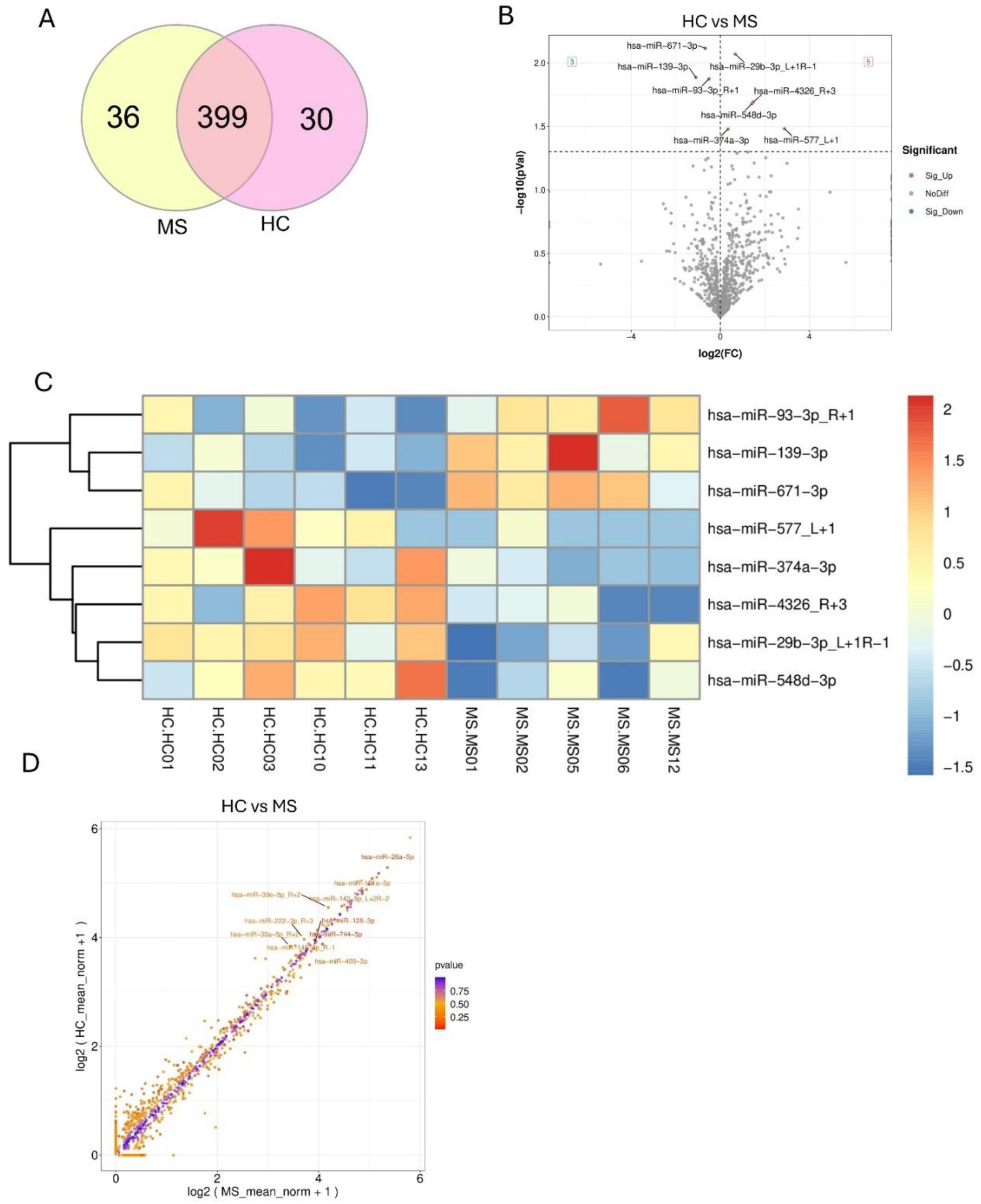
Global profiling of plasma-derived EV-miRNAs reveals distinct signatures in POMS patients compared to matched controls. (A) Venn diagram illustrating shared and unique EV-miRNAs in POMS patients and controls. A total of 465 miRNAs were identified; 399 were shared, 36 were unique to the MS group, and 30 were exclusive to controls. (**B**) Volcano plot depicting differential expression of EV-miRNAs between MS and controls. Significantly upregulated miRNAs in MS are highlighted in red, while significantly downregulated miRNAs are shown in blue (cutoff criteria: |log₂ fold change| > 1.2 and p ≤ 0.05). (**C**) Heatmap demonstrating hierarchical clustering of the top differentially expressed EV-miRNAs across individual MS patients and control samples. Clustering was conducted using Euclidean distance. Expression levels are indicated in red (high expression) and blue (low expression), relative to median expression levels. (**D**) Scatter plot correlation analysis of EV-miRNA expression levels between MS and control samples. Most miRNAs exhibit similar expression patterns across groups; however, certain miRNAs (e.g., miR-26a-5p, miR-142-5p, miR-128-3p) show marked differences.

### Dysregulation of neuroinflammatory, immune, and metabolic pathways in POMS compared to matched controls

To identify biological pathways associated with differentially expressed EV-miRNAs in MS, functional enrichment analyses were conducted using Kyoto Encyclopedia of Genes and Genomes (KEGG) and Gene Ontology (GO) databases. KEGG pathway enrichment analysis (**Figure 2a**) revealed pathways involved in axon guidance (*P* = 2.08 × 10⁻¹⁴, *Q* = 3.50 × 10⁻¹²), PI3K-Akt signaling (*P* = 1.41 × 10⁻¹⁰, *Q* = 9.47 × 10⁻⁹), Rap1 signaling (*P* = 6.35 × 10⁻¹⁰, *Q* = 3.05 × 10⁻⁸), actin cytoskeleton regulation (*P* = 4.36 × 10⁻⁹, *Q* = 1.47 × 10⁻⁷), calcium signaling (*P* = 8.88 × 10⁻⁹, *Q* = 2.71 × 10⁻⁷), ErbB signaling, MAPK signaling, and neurotrophin signaling (**Figure 2aB, 2aC**).

**Figure 2a.**
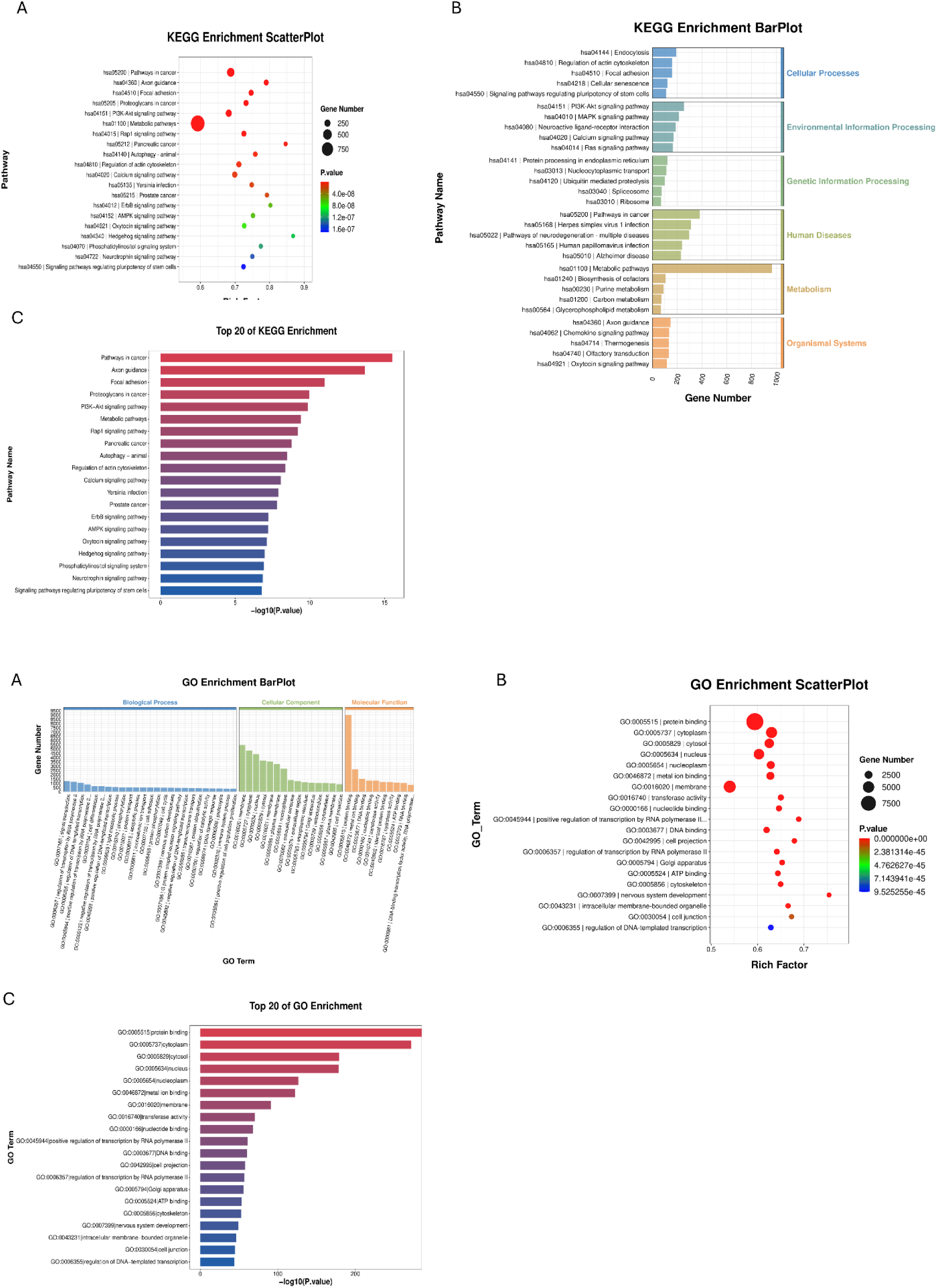
KEGG pathway enrichment analysis of differentially expressed EV-miRNAs in POMS reveals disruption of immune, metabolic, and neurologic signaling. (**A**) KEGG enrichment scatter plot of the top 20 significantly enriched pathways targeted by dysregulated EV-miRNAs. Pathways are arranged by rich factor and color-coded by *P*-value. Circle size represents the number of genes involved per pathway. Enriched pathways include axon guidance, PI3K-Akt signaling, and Rap1 signaling. (**B**) KEGG bar plot categorizing enriched pathways by major biological themes. Categories include metabolism, environmental information processing, human diseases, and organismal systems, highlighting metabolic pathways, MAPK signaling, and neuroactive ligand-receptor interaction. (**C**) Bar graph of the top 20 KEGG pathways ranked by –log₁₀(*P*-value). The most significantly enriched pathways include pathways in cancer, axon guidance, focal adhesion, and PI3K-Akt signaling. **(2b) GO enrichment analysis of target genes of differentially expressed EV miRNAs in POMS highlights perturbations in immune, metabolic, and neurologic processes.** (**A**) GO bar plot summarizing the top enriched GO terms across biological process (blue), cellular component (green), and molecular function (orange) categories. Biological processes include synaptic organization, neuron projection, and regulation of transcription. Cellular components include synapse, nucleus, cytosol, and intracellular organelles. Molecular functions include protein binding, metal ion binding, and nucleotide binding. (**B**) GO enrichment scatter plot of significantly enriched GO terms based on gene ratio and rich factor. Each circle represents a GO term; circle size indicates the number of target genes, and color represents adjusted *P*-value. Processes include protein binding (GO:0005515), cytoskeletal organization, DNA binding, and transcriptional regulation. (**C**) Bar graph of the top 20 GO terms ranked by significance (– log₁₀(*P*-value)). Terms include protein binding, cytoplasmic vesicle composition, transcriptional activity of RNA polymerase II, nervous system development, and regulation of DNA-templated transcription.

GO biological analysis (**Figure 2b**) identified processes such as synaptic function (*P* = 3.00 × 10⁻⁴⁰, *Q* = 1.89 × 10⁻³⁷), neuron projection (*P* = 3.70 × 10⁻³³, *Q* = 1.70 × 10⁻³⁰), neuronal cell body function (*P* = 7.73 × 10⁻³¹, *Q* = 3.06 × 10⁻²⁸), and glutamatergic synapse-related genes (*P* = 3.76 × 10⁻²⁹, *Q* = 1.33 × 10⁻²⁶). Lipid metabolic processes (*P* = 6.77 × 10⁻¹⁹, *Q* = 1.40 × 10⁻¹⁶) were also significantly enriched. Enriched cellular component terms included synapses, axon projections, neuronal junctions, mitochondria, intracellular membrane-bound organelles, Golgi apparatus, and cytoplasmic vesicles. Molecular function analysis highlighted protein binding, metal ion interactions, and RNA polymerase regulatory activity (**Figure 2bB, 2bC**).

### POMS patient EV-miRNA alterations reflect immune and metabolic dysfunction, independent of obesity

To investigate EV-miRNA expression changes specifically associated with POMS independent of obesity, we performed differential expression analysis between healthy-weight POMS patients (MS-NI) and matched healthy-weight controls (HC-NI). Given the exploratory nature of this study and the small sample size, we applied a less stringent statistical threshold (*p* < 0.1) to define significant miRNAs (Supplementary Table 2), while also evaluating miRNAs at more conventional thresholds of *p* < 0.001, *p* < 0.05 and *p* < 0.01 (Table 3). A total of 548 EV-miRNAs were identified, among which miR-5010-3p_R+1, miR-576-3p_R-1, miR-5683, miR-550a-3p, miR-1268b_R-2, miR-1268a, and miR-145-5p were significantly upregulated, whereas miR-1287-5p_R+1, miR-140-3p_R+3, miR-221-3p, miR-10527-5p, miR-181d-5p_R+1, miR-30d-5p_R+2, and miR-361-3p were significantly downregulated in MS-NI compared to HC-NI (Table 3). Out of the 548 miRNAs detected, 465 miRNAs were common to both groups, while 47 were uniquely detected in MS-NI patients and 36 miRNAs were exclusive to HC-NI (**Figure 3A).** Volcano plot analysis provided a global overview of the miRNAs significantly upregulated (red) and downregulated (blue) in MS-NI versus HC-NI samples, highlighting disease-specific EV-miRNA profiles (**Figure 3B).**

**Figure 3.**
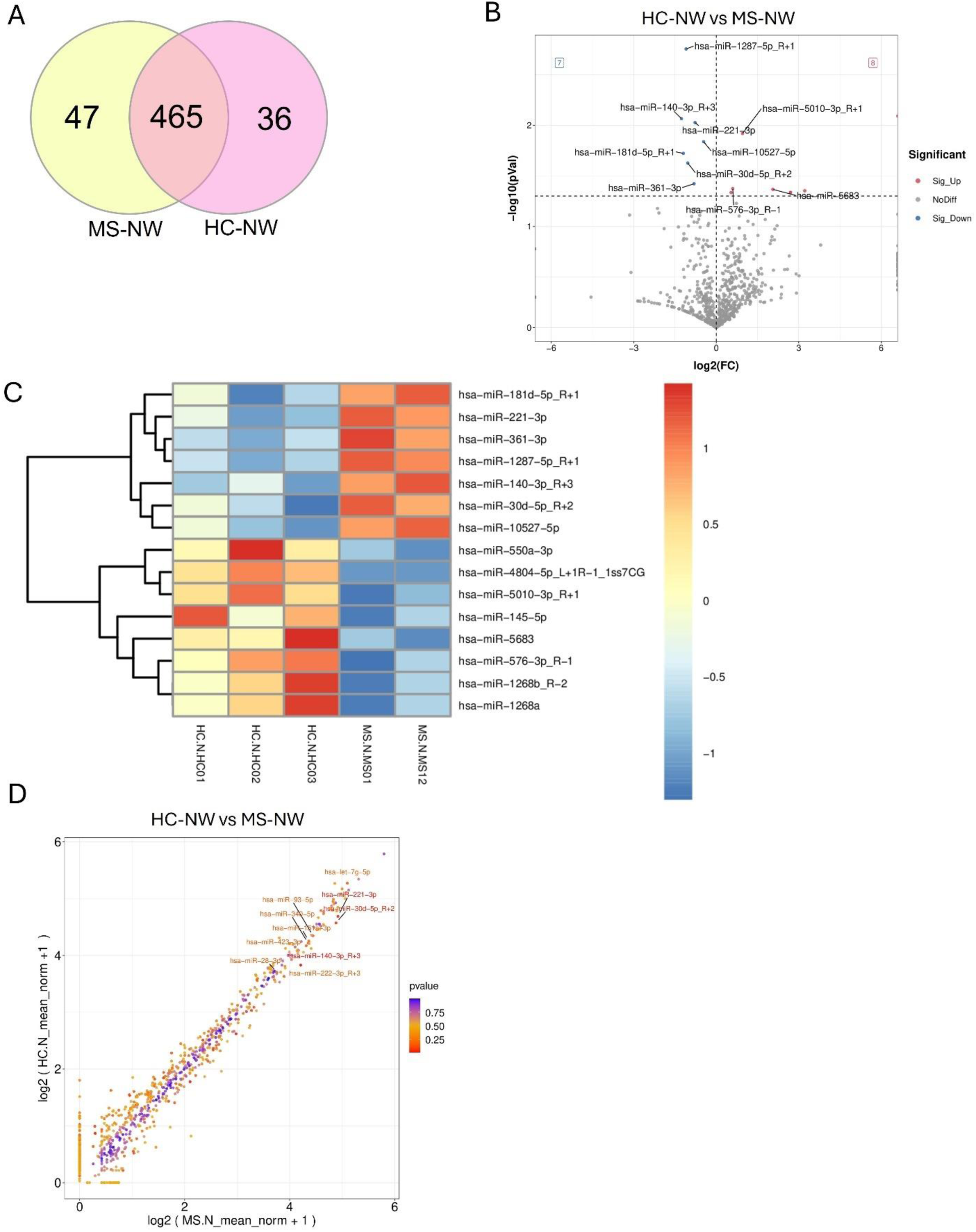
Differential expression of EV-miRNAs in POMS patients reveals disease-specific immune and metabolic regulatory miRNomes. (A) Venn diagram illustrating shared and unique EV-miRNAs in MS-NI and HC-NI. Of 548 identified miRNAs, 465 were common to both groups, 47 unique to MS-NI, and 36 exclusive to HC-NI. (B) Volcano plot depicting differential expression of EV-miRNAs between MS-NI and HC-NI. Red dots represent significantly upregulated miRNAs in MS-NI, while blue dots indicate significantly downregulated miRNAs (cutoff criteria: |log₂ fold change| ≥ 1 and p ≤ 0.05). (C) Heatmap illustrating hierarchical clustering of significantly differentially expressed EV-miRNAs across individual MS-NI and control-NI samples. Clustering utilized Euclidean distance, with red representing high expression and blue indicating low expression relative to median expression levels. (D) Scatter plot correlation analysis comparing EV-miRNA expression levels between MS-NI and control-NI samples, highlighting specific miRNAs such as miR-140-3p_R+3, miR-221-3p, and let-7g-5p with notable differential expression.

**Table 3:** Top differentially expressed EV-miRNAs in HC-NI vs. MS-NI.

| Downregulated DEMs | FC | log2(FC) | p-value* |
| --- | --- | --- | --- |
| miR-1287-5p_R+1 | 0.47 | -1.10 | 1.76E-03 |
| miR-140-3p_R+3 | 0.42 | -1.27 | 8.62E-03 |
| miR-221-3p | 0.59 | -0.77 | 9.41E-03 |
| miR-10527-5p | 0.73 | -0.46 | 1.45E-02 |
| miR-181d-5p_R+1 | 0.44 | -1.20 | 1.90E-02 |
| miR-30d-5p_R+2 | 0.49 | -1.04 | 2.36E-02 |
| miR-361-3p | 0.57 | -0.81 | 3.79E-02 |
| Upregulated DEMs |  |  |  |
| miR-5010-3p_R+1 | 1.94 | 0.96 | 1.21E-02 |
| miR-576-3p_R-1 | 1.52 | 0.60 | 4.23E-02 |
| miR-5683 | 4.17 | 2.06 | 4.30E-02 |
| miR-550a-3p | 9.31 | 3.22 | 4.44E-02 |
| miR-1268b_R-2 | 6.49 | 2.70 | 4.61E-02 |
| **miR-1268a** | 6.49 | 2.70 | 4.61E-02 |
| **miR-145-5p** | 1.45 | 0.54 | 4.63E-02 |

DEM, Differentially expressed miRNAs; FC, fold change; HC-NI, healthy-weight pediatric control; MS-NI, healthy-weight pediatric-onset multiple sclerosis

Hierarchical clustering analysis of differentially expressed miRNAs demonstrated clear segregation between MS-NI and HC-NI samples, suggesting distinct miRNA expression signatures associated with POMS independent of obesity (**Figure 3C).** Correlation analysis indicated consistent miRNA expression patterns for most miRNAs between both groups; however, miRNAs including miR-140-3p_R+3, miR-221-3p, and let-7g-5p showed significant deviations, highlighting specific miRNAs potentially involved in MS pathobiology (**Figure 3D).**

### Functional enrichment analysis of differentially expressed EV-miRNAs in healthy-weight POMS compared to matched, healthy-weight controls

To explore the biological implications of EV-miRNA alterations specifically associated with POMS independent of obesity, functional enrichment analyses using KEGG and GO databases were conducted. KEGG pathway enrichment analysis (**Figure 4a)** identified significant enrichment of pathways involved in axon guidance (P = 1.13 × 10⁻¹², Q = 1.91 × 10⁻¹⁰), Ras signaling (P = 5.96 × 10⁻¹⁰, Q = 5.03 × 10⁻⁸), ErbB signaling (P = 3.73 × 10⁻⁹, Q = 1.72 × 10⁻⁷), Rap1 signaling (P = 1.73 × 10⁻⁸, Q = 5.32 × 10⁻⁷), actin cytoskeleton regulation (P = 4.80 × 10⁻⁸, Q = 1.24 × 10⁻⁶), neurotrophin signaling (P = 2.62 × 10⁻⁷, Q = 5.24 × 10⁻⁶), MAPK signaling (P = 1.05 × 10⁻⁶, Q = 1.49 × 10⁻⁵), calcium signaling (P = 1.63 × 10⁻⁶, Q = 1.90 × 10⁻⁵), and PI3K-Akt signaling (P = 4.22 × 10⁻⁶, Q = 3.98 × 10⁻⁵) (**Figure 4aA–C).**

**Figure 4a.**
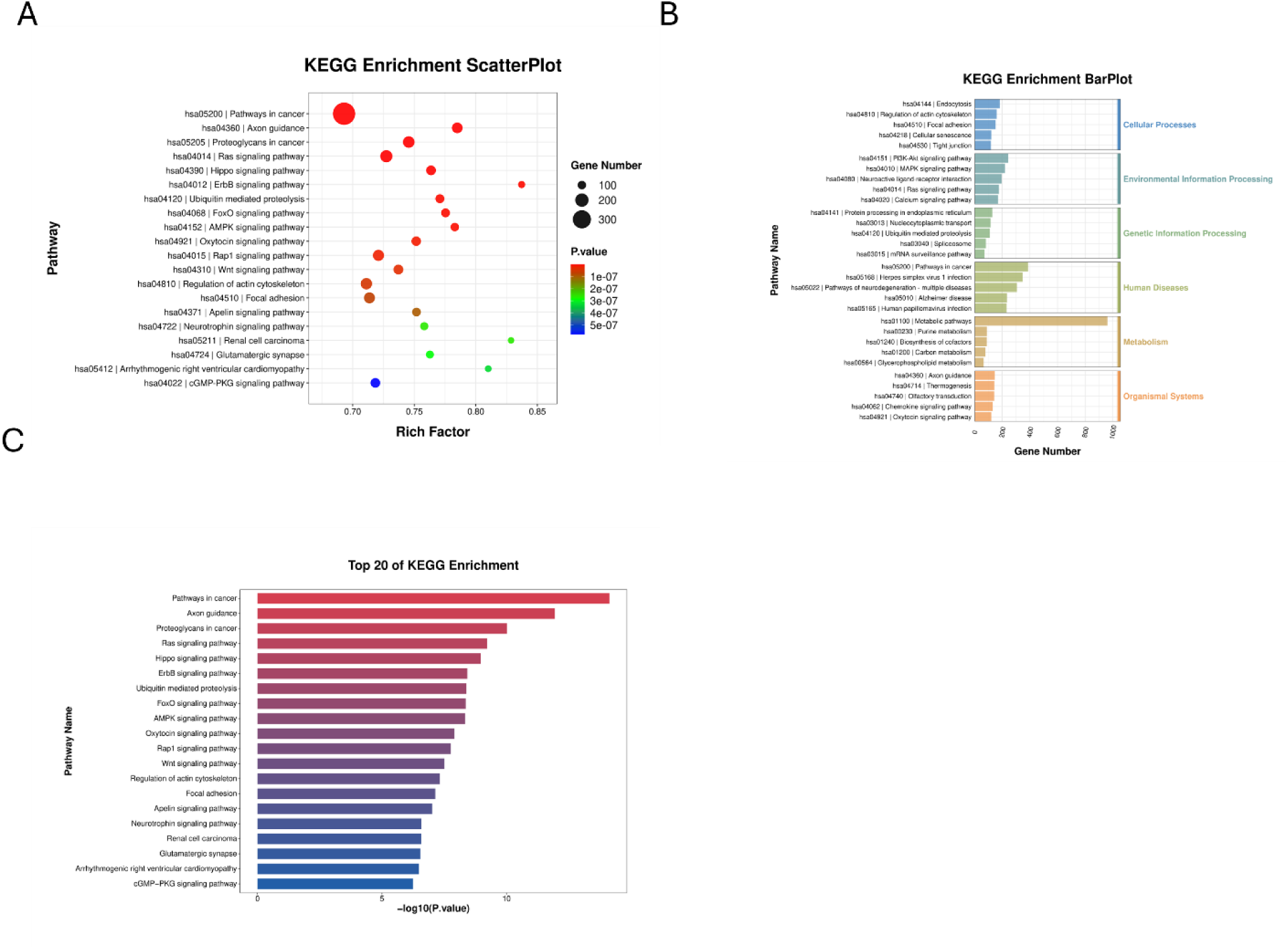
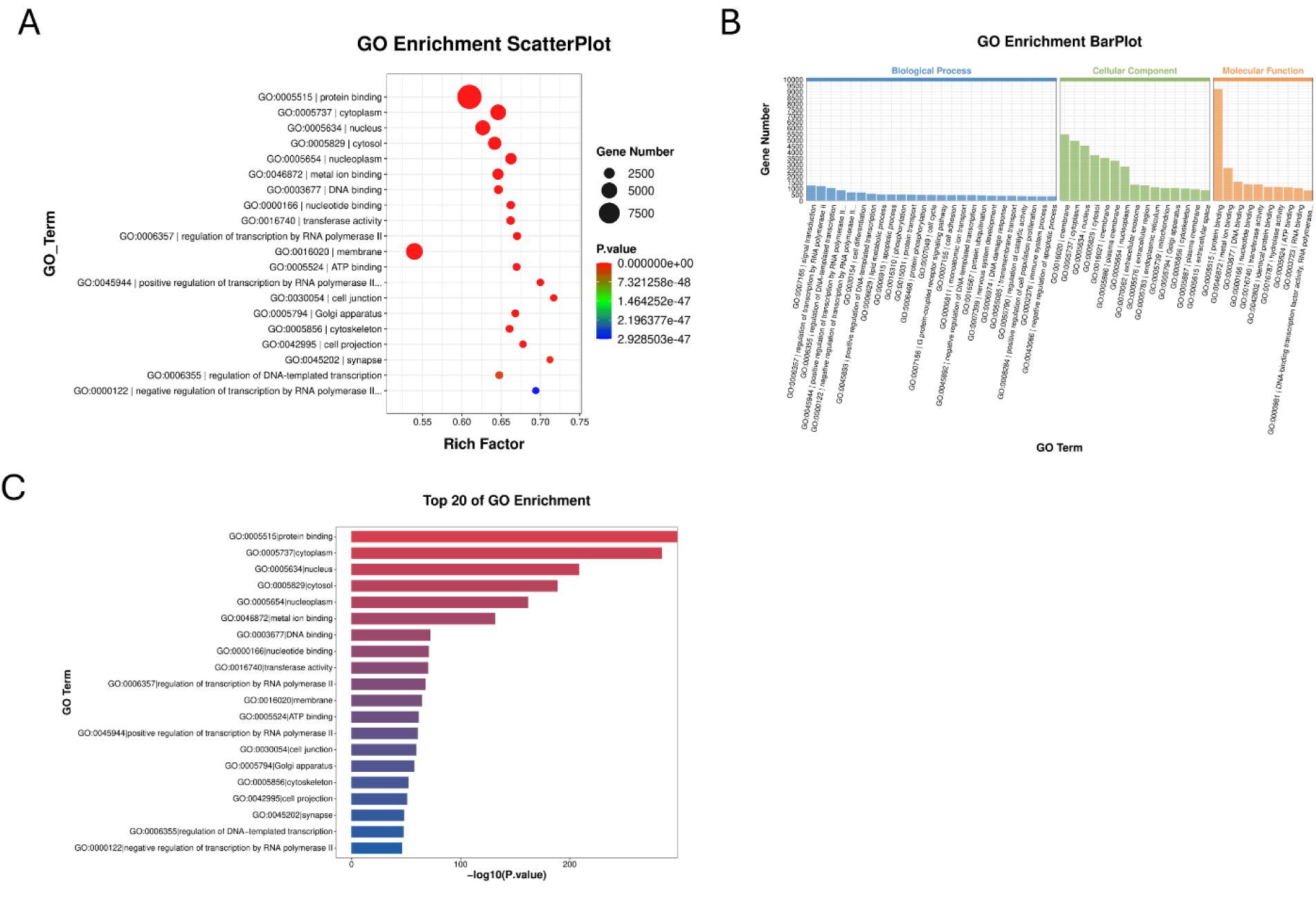
KEGG pathway enrichment analysis of differentially expressed EV-miRNAs in MS-NI versus controls (PC-NI) reveals disrupted immune, metabolic, and neuronal signaling. (A) KEGG enrichment scatter plot illustrating significantly enriched pathways targeted by dysregulated EV-miRNAs. Circle size corresponds to the number of target genes, and color represents adjusted P-value significance. Pathways include axon guidance, Ras signaling, ErbB signaling, Rap1 signaling, and PI3K-Akt signaling. (B) KEGG enrichment bar plot categorizing pathways by biological function, including environmental information processing, metabolism, neurodevelopment, and immune responses. Notable pathways include MAPK signaling, calcium signaling, and actin cytoskeleton regulation. (C) Bar graph depicting the top 20 KEGG pathways ranked by significance (–log₁₀(P-value)), emphasizing axon guidance, Rap1 signaling, ErbB signaling, focal adhesion, and neurotrophin signaling.**Figure 4b. GO enrichment analysis of EV-miRNA targets in MS-NI versus matched HC-NI identifies alterations in neuronal signaling, lipid metabolism, and immune regulation.** (A) GO enrichment scatter plot presenting significantly enriched terms across biological processes, cellular components, and molecular functions. Circle size denotes gene count, and color indicates adjusted P-value significance. Prominent terms include protein binding, cytoplasm, nucleus, metal ion binding, and DNA binding. (B) GO enrichment bar plot summarizing significantly enriched terms across GO categories. Highlighted biological processes include synaptic function, neuron projection, and immune signaling. Cellular components comprise synapse, cytoskeleton, Golgi apparatus, and mitochondrion. Molecular functions include lipid binding, nucleotide binding, and transferase activity. (C) Bar plot illustrating the top 20 GO terms ranked by significance (–log₁₀(P-value)), with major terms including glutamatergic synapse, axon, lipid metabolism, DNA-binding transcription factor activity, and mitochondrial matrix.

GO biological process analysis (Figure 4b) highlighted significantly enriched terms such as synaptic function (P = 4.51 × 10⁻⁴⁹, Q = 4.28 × 10⁻⁴⁶), glutamatergic synapse regulation (P = 3.79 × 10⁻³⁴, Q = 1.85 × 10⁻³¹), neuron projection (P = 5.07 × 10⁻²⁹, Q = 1.88 × 10⁻²⁶), axon-related processes (P = 4.33 × 10⁻²⁸, Q = 1.51 × 10⁻²⁵), neuronal cell body regulation (P = 5.05 × 10⁻²⁴, Q = 1.49 × 10⁻²¹), lipid metabolic processes (P = 2.26 × 10⁻¹³, Q = 2.75 × 10⁻¹¹), and lipid binding (P = 2.48 × 10⁻¹², Q = 2.70 × 10⁻¹⁰) (**Figure 4bA, B**).

GO cellular component analysis revealed significant enrichment of terms related to synaptic vesicles, axonal growth processes, cytoskeletal structures, mitochondrial components, Golgi apparatus, and intracellular membrane-bounded organelles. GO molecular function analysis emphasized protein binding, metal ion binding, nucleotide binding, lipid binding, DNA-binding transcription factor activity, and transferase activity (**Figure 4bB, C**).

### EV-miRNA profiling in obese multiple sclerosis (POMS-Ob) versus matched controls (PC-Ob) reveals metabolic and inflammatory disturbances

To identify EV-miRNA expression changes specifically associated with POMS in an obese context, differential expression analysis was conducted comparing POMS-Ob and matched PC-Ob. Given the exploratory nature of this study and the small sample size, we applied a less stringent statistical threshold (*p* < 0.1) to define significant miRNAs (Supplementary Table 3), while also evaluating miRNAs at more conventional thresholds of *p* < 0.001, *p* < 0.05 and *p* < 0.01 (Table 4). A total of 520 EV-miRNAs were detected; miR-142-5p, miR-29a-3p, and miR-103a-2-5p were significantly upregulated, while miR-23a-3p, miR-671-3p, and miR-20a-3p exhibited marked downregulation in the POMS-Ob group compared to PC-Ob (**Table 4).** Among these miRNAs, 420 were shared by both groups, 36 were exclusive to POMS-Ob, and 64 were unique to PC-Ob (**Figure 5A).** Volcano plot analysis provided an overview of significantly upregulated (red) and downregulated (blue) EV-miRNAs in POMS-Ob versus PC-Ob samples, highlighting distinct EV-miRNA profiles linked to POMS in obese patients (**Figure 5B).**

**Figure 5.**
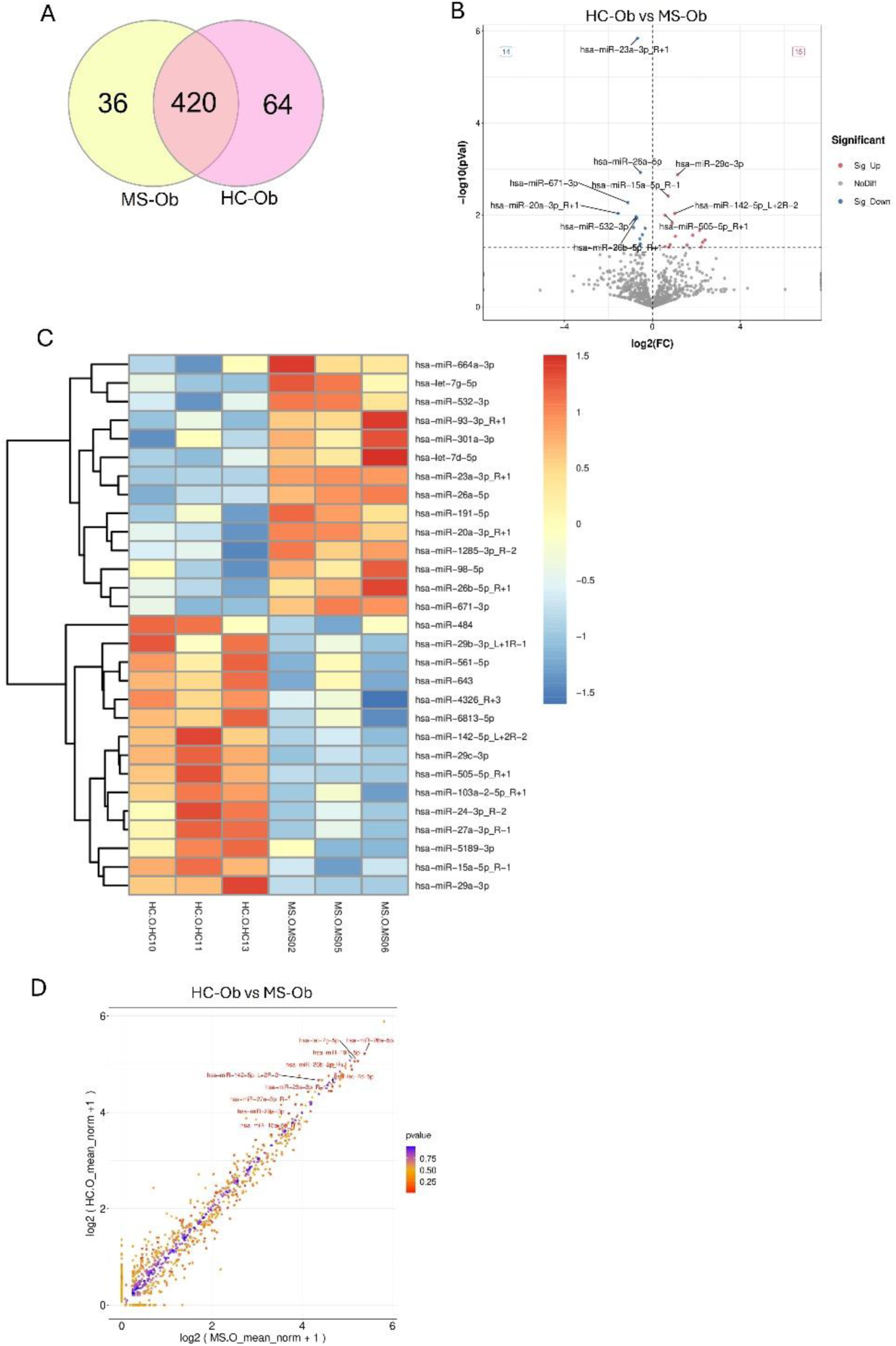
EV-miRNA expression profiles in obese POMS patients compared to obese controls. (A) Venn diagram showing distribution of EV-miRNAs in POMS-Ob and PC-Ob. Out of 520 miRNAs detected, 420 were common, 36 unique to POMS-Ob, and 64 exclusive to PC-Ob. (B) Volcano plot representing differential expression of EV-miRNAs between POMS-Ob and PC-Ob. Significantly upregulated miRNAs are highlighted in red, significantly downregulated in blue (cutoff criteria: |log₂ fold change| ≥ 1, p ≤ 0.05). (C) Heatmap illustrating hierarchical clustering of significantly differentially expressed EV-miRNAs between POMS-Ob and PC-Ob samples. (D) Scatter plot correlation analysis comparing EV-miRNA expression between POMS-Ob and PC-Ob groups, highlighting miRNAs with notable differential expression.

**Table 4:** Top differentially expressed EV-miRNAs in PC-Ob vs. POMS-Ob.

| Downregulated DEMs | FC | log<sub>2</sub>(FC) | p-value* |
| --- | --- | --- | --- |
| miR-23a-3p_R+1 | 0.63 | -0.67 | 1.43E-06 |
| miR-26a-5p | 0.69 | -0.54 | 1.19E-03 |
| miR-671-3p | 0.46 | -1.11 | 5.40E-03 |
| miR-20a-3p_R+1 | 0.34 | -1.55 | 9.27E-03 |
| miR-532-3p | 0.6 | -0.75 | 1.09E-02 |
| miR-26b-5p_R+1 | 0.61 | -0.7 | 1.18E-02 |
| miR-93-3p_R+1 | 0.57 | -0.81 | 1.27E-02 |
| miR-1285-3p_R-2 | 0.55 | -0.85 | 1.88E-02 |
| miR-191-5p | 0.79 | -0.33 | 1.96E-02 |
| let-7d-5p | 0.73 | -0.46 | 2.70E-02 |
| let-7g-5p | 0.68 | -0.57 | 3.31E-02 |
| miR-98-5p | 0.68 | -0.55 | 4.27E-02 |
| miR-301a-3p | 0.67 | -0.58 | 4.53E-02 |
| miR-664a-3p | 0.56 | -0.84 | 4.96E-02 |
| **Upregulated DEMs** |  |  |  |
| miR-29c-3p | 2.22 | 1.15 | 1.34E-03 |
| miR-15a-5p_R-1 | 1.65 | 0.72 | 3.90E-03 |
| miR-142-5p_L+2R-2 | 2.02 | 1.01 | 9.33E-03 |
| miR-505-5p_R+1 | 1.49 | 0.58 | 1.01E-02 |
| miR-29a-3p | 1.87 | 0.9 | 1.50E-02 |
| miR-103a-2-5p_R+1 | 4.43 | 2.15 | 2.16E-02 |
| miR-6813-5p | 3.57 | 1.84 | 2.74E-02 |
| miR-27a-3p_R-1 | 2.06 | 1.05 | 2.90E-02 |
| miR-5189-3p | 5.26 | 2.4 | 3.51E-02 |
| miR-561-5p | 4.9 | 2.29 | 3.93E-02 |
| miR-24-3p_R-2 | 1.75 | 0.81 | 4.44E-02 |
| miR-4326_R+3 | 2.99 | 1.58 | 4.54E-02 |
| miR-484 | 1.49 | 0.58 | 4.86E-02 |
| miR-643 | 4.67 | 2.22 | 4.96E-02 |
| miR-29b-3p_L+1R-1 | 1.7 | 0.76 | 4.99E-02 |

DEM, Differentially expressed miRNAs; FC, fold change; PC-Ob, obese control; POMS-Ob, obese pediatric-onset multiple sclerosis

Hierarchical clustering analysis of differentially expressed miRNAs demonstrated distinct expression signatures differentiating POMS-Ob from PC-Ob groups, reflecting potential metabolic and inflammatory disturbances associated with POMS in obesity (**Figure 5C).** Correlation analysis showed consistent miRNA expression patterns for most miRNAs; however, miR-142-5p, miR-29a-3p, and let-7d-5p exhibited notably elevated expression, while miR-15a-5p, miR-671-3p, and miR-505-5p demonstrated significant downregulation in POMS-Ob patients compared to PC-Ob (**Figure 5D).**

### Functional enrichment analysis of differentially expressed miRNAs in obese POMS vs. obese controls reveals immune, metabolic, and neurodegenerative

To elucidate the biological pathways associated with altered EV-miRNAs in POMS-Ob, functional enrichment analyses using KEGG and GO databases were performed. KEGG pathway enrichment analysis (**Figure 6a)** identified significantly enriched pathways including axon guidance (P = 6.66 × 10⁻¹³, Q = 5.61 × 10⁻¹¹), calcium signaling (P = 1.46 × 10⁻¹², Q = 9.85 × 10⁻¹¹), focal adhesion (P = 1.31 × 10⁻⁹, Q = 4.42 × 10⁻⁸), PI3K-Akt signaling (P = 6.21 × 10⁻⁹, Q = 1.74 × 10⁻⁷), MAPK signaling (P = 2.46 × 10⁻⁷, Q = 4.39 × 10⁻⁶), and neurotrophin signaling (P = 4.04 × 10⁻⁶, Q = 3.58 × 10⁻⁵) **(Figure 6aA–C**).

**Figure 6a.**
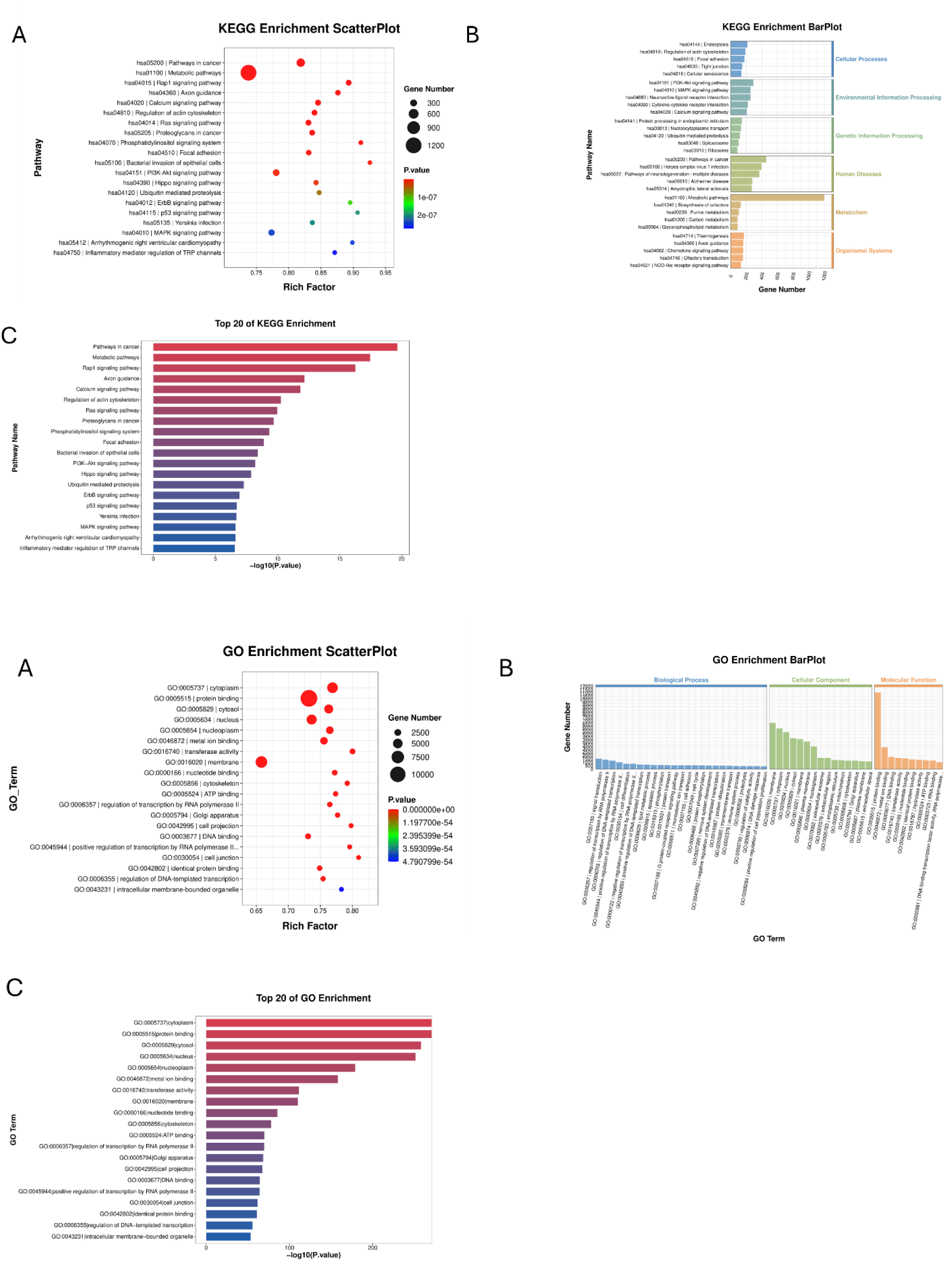
KEGG enrichment analysis of EV-miRNA targets in obese POMS patients versus obese controls reveals activation of neuroimmune and metabolic signaling pathways. (A) KEGG enrichment scatter plot illustrating significantly enriched pathways targeted by dysregulated EV-miRNAs. Circle size corresponds to gene count, and color represents adjusted P-value significance. Pathways prominently enriched include axon guidance, calcium signaling, focal adhesion, Rap1 signaling, and PI3K-Akt signaling. (B) KEGG bar plot categorizing pathways by major biological functions, including metabolism, environmental information processing, human diseases, and organismal systems. Highlighted pathways include MAPK signaling, PI3K-Akt signaling, and neuroactive ligand-receptor interaction. (C) Bar graph showing the top 20 KEGG pathways ranked by significance (–log₁₀(P-value)). The most significantly enriched pathways include focal adhesion, phosphatidylinositol signaling, Hippo signaling, and TRP channel regulation. **(6b) GO enrichment analysis of differentially expressed EV-miRNA targets in obese MS patients identifies disruptions in synaptic structure, lipid metabolism, and immune regulatory functions.** (A) GO enrichment scatter plot presenting significantly enriched terms across biological processes, cellular components, and molecular functions. Circle size indicates the number of genes involved; color represents adjusted P-value significance. Key terms include cytoplasm, protein binding, nucleoplasm, membrane components, and metal ion binding. (B) GO enrichment bar plot summarizing highly enriched terms across major GO categories. Highlighted biological processes include neuron projection, synaptic development, immune signaling, and lipid metabolism. Cellular component terms feature synapse, Golgi apparatus, and mitochondrion. Molecular function terms emphasize nucleotide binding, kinase activity, and transferase activity. (C) Bar plot illustrating the top 20 GO terms ranked by significance (–log₁₀(P-value)), highlighting cytoplasm, protein binding, lipid transport, axon development, and mitochondrial structures.

GO biological process enrichment analysis (**Figure 6b)** revealed significantly enriched terms including neuron projection (P = 3.46 × 10⁻³⁷, Q = 1.35 × 10⁻³⁴), neuronal cell body function (P = 4.57 × 10⁻³⁶, Q = 1.71 × 10⁻³³), axon-related structures (P = 7.67 × 10⁻³⁰, Q = 2.23 × 10⁻²⁷), lipid metabolic processes (P = 2.28 × 10⁻²⁵, Q = 5.48 × 10⁻²³), and postsynaptic density (P = 5.28 × 10⁻²⁴, Q = 1.16 × 10⁻²¹) (**Figure 6bA, B**). Additionally, cytokine signaling, antigen presentation, T-cell activation, cholesterol metabolism, oxidative stress regulation, and synaptic transmission processes were significantly enriched.

GO cellular component enrichment analysis highlighted significant enrichment in protein-containing complexes, synaptic vesicles, mitochondrial structures, Golgi apparatus, nucleoplasm, membrane components, and cytoplasmic structures. GO molecular function analysis underscored protein binding, metal ion binding, nucleotide binding, lipid binding, DNA-binding transcription factor activity, and transferase activity (**Figure 6bC**).

### EV-miRNA alterations in obese multiple sclerosis patients highlight metabolic and neuroimmune dysregulation

To determine the influence of obesity on EV-miRNA profiles in POMS, plasma EV-miRNA profiles were compared between POMS-Ob and MS-Nl. Given the exploratory nature of this study and the small sample size, we applied a less stringent statistical threshold (*p* < 0.1) to define significant miRNAs (Supplementary Table 4), while also evaluating miRNAs at more conventional thresholds of *p* < 0.001, *p* < 0.05 and *p* < 0.01 (Table 5). A total of 533 miRNAs were detected; notably, miR-140-3p_R+3, miR-30d-5p_R+2, miR-296-5p, mir-1302-1-p5_1ss9AG, and miR-29b-1-5p_R-1 exhibited significant upregulation (fold change > 3.0 or infinite), whereas miR-27a-5p showed marked downregulation (fold change = 0.41) in POMS-Ob relative to MS-NI (**Table 5).** Out of these miRNAs, 435 were common to both groups, 21 were exclusive to POMS-Ob, and 77 were uniquely detected in MS-NI (Figure 7A). Volcano plot analysis highlighted significantly upregulated (red) and downregulated (blue) EV-miRNAs distinguishing POMS-Ob from MS-NI, reflecting distinct EV-miRNA signatures related to obesity in POMS (**Figure 7B).**

**Figure 7.**
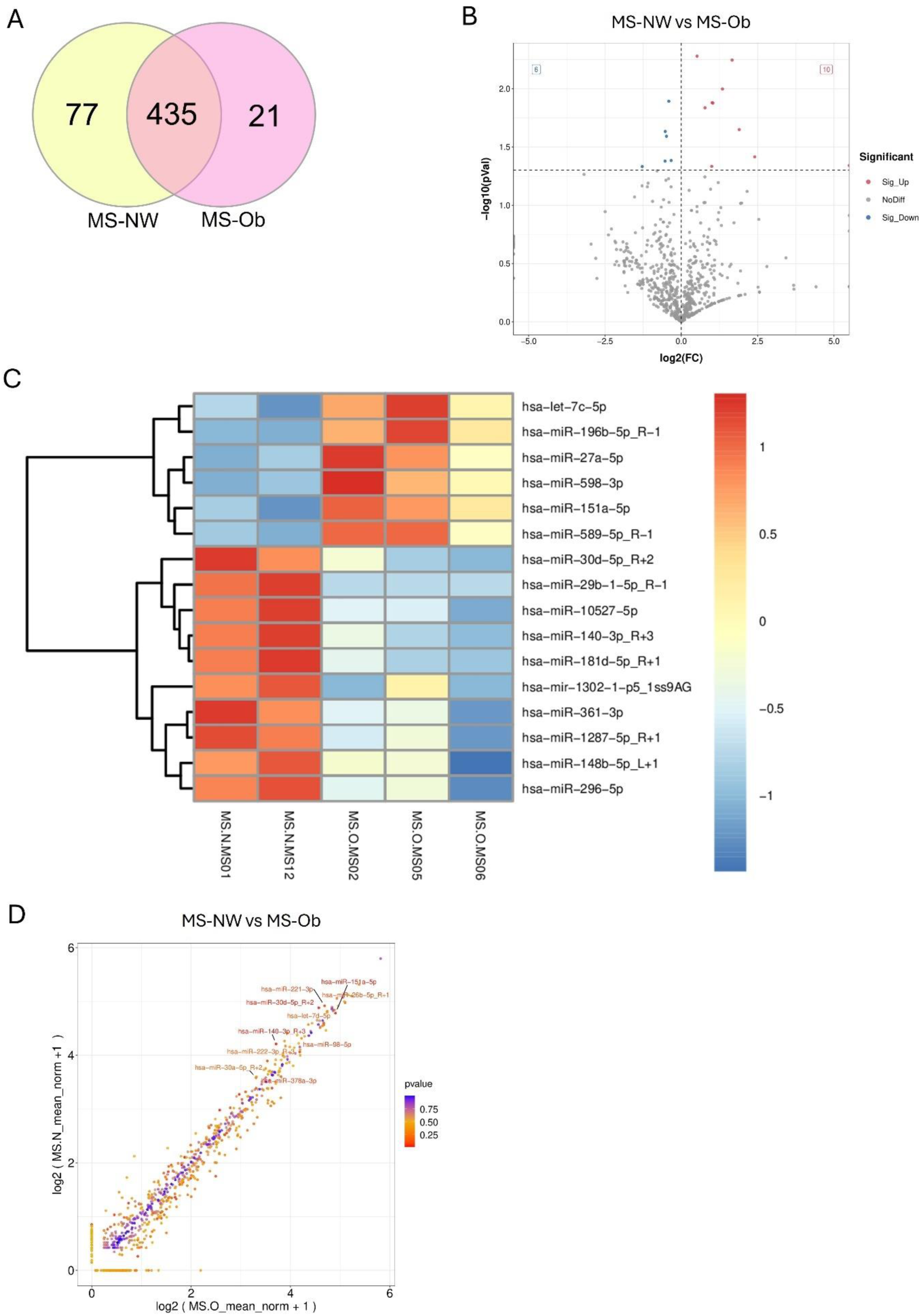
EV-miRNA expression profiles in obese POMS-Ob compared to non-obese POMS-NI. (A) Venn diagram illustrating EV-miRNA distribution identified in POMS-Ob and MS-NI groups. Among 533 miRNAs detected, 435 were shared, 21 unique to POMS-Ob, and 77 exclusive to MS-NI. (B) Volcano plot representing differential EV-miRNA expression between POMS-Ob and MS-NI. Significantly upregulated miRNAs are highlighted in red and significantly downregulated miRNAs in blue (cutoff criteria: |log₂ fold change| ≥ 1, p ≤ 0.05). (C) Heatmap depicting hierarchical clustering of significantly differentially expressed EV-miRNAs between POMS-Ob and MS-NI groups. (D) Scatter plot correlation analysis comparing EV-miRNA expression between POMS-Ob and MS-NI groups, highlighting specific miRNAs demonstrating significant differential expression.

**Table 5:**
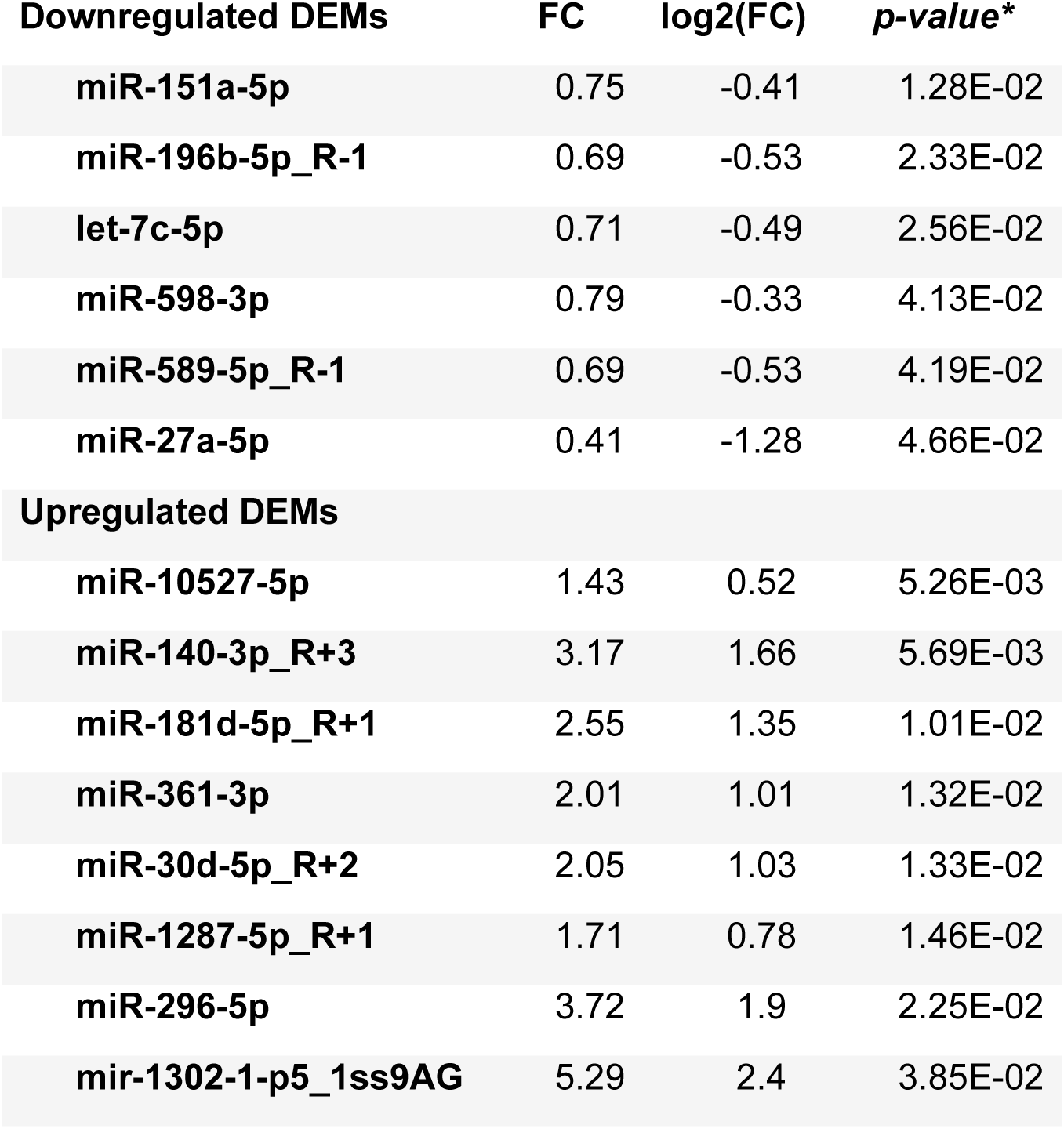
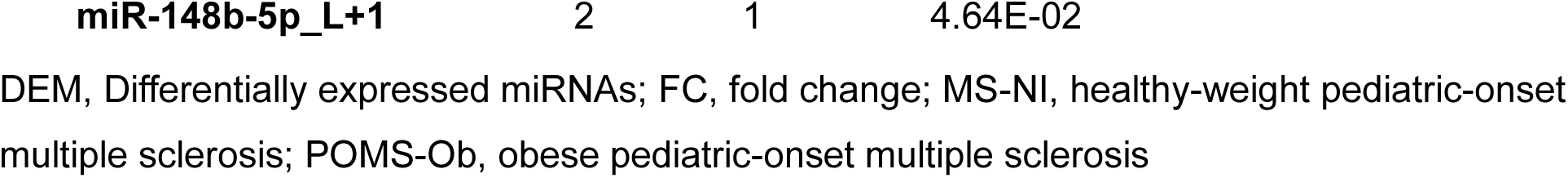
Top differentially expressed EV-miRNAs in MS-NI vs. POMS-Ob.

| Downregulated DEMs | FC | log2(FC) | *p-value* * |
| --- | --- | --- | --- |
| miR-151a-5p | 0.75 | -0.41 | 1.28E-02 |
| miR-196b-5p_R-1 | 0.69 | -0.53 | 2.33E-02 |
| let-7c-5p | 0.71 | -0.49 | 2.56E-02 |
| miR-598-3p | 0.79 | -0.33 | 4.13E-02 |
| miR-589-5p_R-1 | 0.69 | -0.53 | 4.19E-02 |
| miR-27a-5p | 0.41 | -1.28 | 4.66E-02 |
| **Upregulated DEMs** |  |  |  |
| miR-10527-5p | 1.43 | 0.52 | 5.26E-03 |
| miR-140-3p_R+3 | 3.17 | 1.66 | 5.69E-03 |
| miR-181d-5p_R+1 | 2.55 | 1.35 | 1.01E-02 |
| miR-361-3p | 2.01 | 1.01 | 1.32E-02 |
| miR-30d-5p_R+2 | 2.05 | 1.03 | 1.33E-02 |
| miR-1287-5p_R+1 | 1.71 | 0.78 | 1.46E-02 |
| miR-296-5p | 3.72 | 1.9 | 2.25E-02 |
| mir-1302-1-p5_1ss9AG | 5.29 | 2.4 | 3.85E-02 |
| **miR-148b-5p_L+1** | 2 | 1 | 4.64E-02 |

DEM, Differentially expressed miRNAs; FC, fold change; MS-NI, healthy-weight pediatric-onset multiple sclerosis; POMS-Ob, obese pediatric-onset multiple sclerosis

Hierarchical clustering analysis clearly differentiated miRNA expression profiles between POMS-Ob and MS-NI groups, indicating distinct metabolic and neuroimmune dysregulation linked to obesity in POMS patients (**Figure 7C).** Correlation analysis revealed consistent expression for most miRNAs; however, miRNAs including miR-140-3p_R+3, miR-30d-5p_R+2, and let-7c-5p were notably increased, whereas miR-361-3p, miR-378a-3p, and miR-296-5p exhibited decreased expression in POMS-Ob compared to MS-NI (**Figure 7D).**

### Functional enrichment analysis of differentially expressed miRNAs in obese MS vs. non-obese MS identifies metabolic and neuroinflammatory pathway disruptions

To identify functional pathways associated with EV-miRNA alterations induced by obesity in POMS, KEGG and GO enrichment analyses were conducted on miRNAs differentially expressed between POMS-Ob and MS-NI. KEGG pathway enrichment analysis (**Figure 8a)** revealed significant enrichment in PI3K-Akt signaling (P = 5.73 × 10⁻¹⁰, Q = 1.93 × 10⁻⁸), MAPK signaling (P = 9.41 × 10⁻¹¹, Q = 4.96 × 10⁻⁹), cytokine–cytokine receptor interaction, Rap1 signaling (P = 5.41 × 10⁻¹², Q = 6.08 × 10⁻¹⁰), Ras signaling, ErbB signaling, calcium signaling, axon guidance (P = 2.47 × 10⁻¹¹, Q = 2.08 × 10⁻⁹), regulation of actin cytoskeleton (P = 4.05 × 10⁻¹², Q = 6.08 × 10⁻¹⁰), and focal adhesion (**Figure 8aA–C)**.

**Figure 8a.**
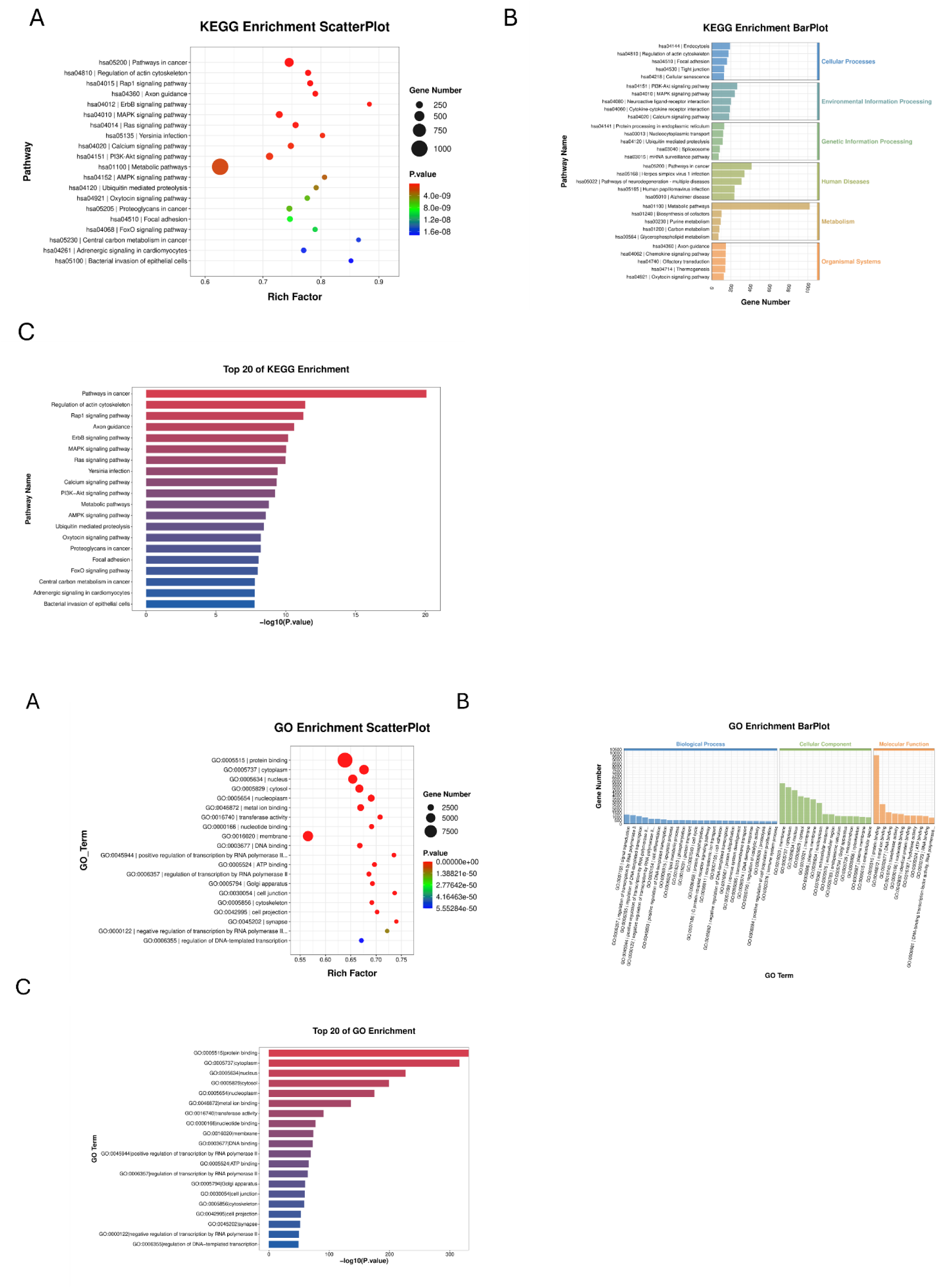
KEGG pathway enrichment analysis of EV-miRNA targets in obese POMS-Ob compared to non-obese POMS-NI. (A) KEGG enrichment scatter plot illustrating significantly enriched pathways targeted by differentially expressed EV-miRNAs. Circle size represents gene count; color indicates adjusted P-value significance. Enriched pathways include PI3K-Akt signaling, MAPK signaling, cytokine–cytokine receptor interaction, Rap1 signaling, axon guidance, ErbB signaling, and calcium signaling. (B) KEGG enrichment bar plot categorizing pathways into major biological domains, including metabolism, immune signaling, and organismal systems. Highlighted pathways include neuroactive ligand-receptor interaction, glycerophospholipid metabolism, and endocytosis. (C) Bar graph of the top 20 KEGG pathways ranked by significance (–log₁₀(P-value)), emphasizing regulation of actin cytoskeleton, Rap1 signaling, focal adhesion, and central carbon metabolism in cancer. **(8b) GO enrichment analysis of predicted targets of dysregulated EV miRNAs in obese MS compared to normal-weight MS reveals altered neuronal structure, lipid metabolism, and intracellular signaling.** (A) GO enrichment scatter plot presenting significantly enriched GO terms across biological processes, cellular components, and molecular functions. Circle size corresponds to gene count; color indicates adjusted P-value significance. Prominent enriched terms include protein binding, cytoplasm, nucleus, metal ion binding, membrane components, and DNA binding. (B) GO enrichment bar plot summarizing highly enriched terms across GO domains. Highlighted biological processes include lipid metabolism, immune signaling, neuron projection, and synaptic organization. Cellular components include synapse, Golgi apparatus, mitochondrion, and intracellular organelles. Molecular functions emphasize nucleotide binding, kinase activity, and transferase activity. (C) Bar plot illustrating the top 20 GO terms ranked by significance (–log₁₀(P-value)), including cytoplasm, protein binding, lipid metabolic processes, and axon development.

GO biological process enrichment analysis (**Figure 8b)** identified significantly enriched terms such as protein binding, lipid metabolic processes, neuron projection, immune signaling, synaptic organization, and glutamatergic synapse. Cellular component enrichment analysis highlighted significant enrichment of cytoplasm (P = 2.08 × 10⁻³¹⁶, Q = 1.80 × 10⁻³¹²), nucleus, cytosol, membrane components, Golgi apparatus, mitochondria, synaptic vesicles, axonal structures, and intracellular membrane-bounded organelles (**Figure 8bA–C)**. Molecular function analysis underscored metal ion binding, nucleotide binding, transferase activity, RNA polymerase regulatory activity, and DNA binding (**Figure 8bB**).

## Discussion

This study provides novel insights into the role of EV-miRNAs in POMS, emphasizing how obesity further modifies these molecular signatures. A critical methodological strength of our approach was the use of age-, sex-, and BMI-matched control samples, ensuring that observed miRNA alterations reflect disease-specific biology rather than confounding demographic variability. Our data reveal that MS-specific EV-miRNA dysregulation is closely linked to neuroinflammation, immune dysregulation, and altered lipid metabolism. Notably, obesity introduces additional perturbations in metabolic pathways, which may provide clues into adiposity-associated CNS autoimmunity. Given miRNAs’ pivotal role in post-transcriptional gene regulation, these findings underscore their potential as both biomarkers and mechanistic agents in MS pathophysiology.

### EV miRNAs as regulators of neuroinflammation and immune responses in MS

One of the key findings of this study was the distinct miRNA expression pattern observed in POMS patients compared to controls, with several miRNAs exhibiting significant dysregulation. Many of these differentially expressed miRNAs are known regulators of immune activation and neuroinflammation, further supporting their role in MS pathogenesis. For example, miR-29b-3p, which was significantly upregulated in MS patients, has been linked to cholesterol metabolism and myelin integrity, suggesting that its altered expression may contribute to demyelination and lipid imbalance in MS[35–38]. Conversely, the downregulation of miR-671-3p and miR-139-3p, which are involved in BBB integrity and immune suppression [39,40], suggests a loss of regulatory control over neurovascular homeostasis, potentially exacerbating CNS inflammation[41–43]. The role of immune system-related pathways, including cytokine–cytokine receptor interactions and T-cell receptor signaling, further supports the hypothesis that EV miRNAs contribute to immune cell activation and inflammatory responses in MS[44–46] For instance, several EV-miRNAs identified in our analysis (such as miR-146a-5p and miR-223-3p) are known to modulate NF-κB activation and macrophage polarization, implicating these miRNAs in orchestrating pro-inflammatory cascades that contribute to MS pathology[47,48] [49,50] .

### Obesity-driven miRNA alterations exacerbate metabolic, lipid metabolic, and neuroinflammatory dysregulation in MS

A significant finding of this study is that obesity amplifies dysregulation in the EV-miRNAs involved in immune, metabolic, and lipid imbalances. Obesity is known to promote chronic low-grade inflammation and increase susceptibility to autoimmune disorders through altered adipokine and cytokine signaling[15,51–54]. Our analysis identified significant alterations in miRNAs related to adipogenesis, inflammation, and lipid metabolism in obese POMS patients, underscoring obesity’s role in promoting MS pathogenesis.

Specifically, miRNAs such as miR-29a-3p, miR-223-3p, let-7c-5p, miR-142-5p, miR-146a-5p, and miR-151a-5p were significantly elevated, suggesting their involvement in inflammatory lipid signaling, cholesterol metabolism, and immune activation [55–63] . Conversely, miRNAs including miR-15a-5p, miR-671-3p, miR-320b, miR-505-5p, and let-7c-5p were notably downregulated, suggesting disruptions in lipid oxidation, energy balance, neurovascular unit integrity, and increased oxidative stress [57–62,64–72].

KEGG pathway analysis further confirmed significant alterations in lipid-processing enzymes, transporters, and regulators. Enriched pathways, such as fatty acid degradation, sphingolipid signaling, glycerophospholipid metabolism, and cholesterol metabolism, indicate their pivotal roles in CNS lipid homeostasis, myelin synthesis, and immune regulation[63,73,74]. GO analysis similarly emphasized significant enrichment in lipid metabolic processes, lipid binding, cholesterol binding, and phospholipid transport, demonstrating miRNAs’ regulatory roles in lipid-mediated neuroinflammation and metabolic homeostasis[75–77]. Disruptions in lipid metabolism are closely associated with enhanced neuroinflammatory responses[78]. Collectively, these obesity-driven EV-miRNA alterations suggest intensified pro-inflammatory lipid signaling and neuroimmune activation, potentially exacerbating CNS inflammation and demyelination via mechanisms involving lipid accumulation in microglia and astrocytes [79] [80].

### Functional implications of enriched pathways: linking immune regulation, metabolism, and neurodegeneration

Our enrichment analyses identified key pathways relevant to MS pathogenesis, notably cytokine–cytokine receptor interactions, T-cell receptor signaling, and NF-κB cascades, underscoring miRNAs’ roles in regulating immune cell activation and inflammatory cytokine production[18,40,44,45,81]. This finding aligns with previous studies demonstrating that miRNAs are crucial modulators of innate and adaptive immune responses in neuroinflammatory disease[59]. For example, miR-223 and miR-146a are involved in negative feedback loops that control immune activation; their dysregulation in MS could lead to excessive inflammation[17,48–50,61].

Additionally, our analyses highlighted enrichment of lipid metabolism pathways, including cholesterol transport and fatty acid oxidation, indicating that miRNA dysregulation also contributes to metabolic dysfunction in MS[28,69,76]. Myelin, a lipid-rich structure, can be compromised by altered lipid metabolism, directly impacting remyelination and contributing to neurodegeneration[63,73,82,83]. The observed dysregulation of lipid-associated miRNAs, such as elevated miR-29 and reduced let-7 families, may further impair myelin maintenance, remyelination, and neuronal integrity[38,55]. Such metabolic alterations potentially amplify inflammation, thereby exacerbating neuronal injury and cognitive deficits, which is consistent with clinical data linking obesity to MS progression[73,84].

These insights collectively highlight the intersection of immune dysregulation, altered lipid metabolism, and neurodegeneration, driven by EV-miRNA-mediated pathways in POMS, particularly in the context of obesity.

### Clinical relevance and future directions

The identification of disease- and obesity-specific EV-miRNAs highlights their potential as minimally invasive biomarkers for MS diagnosis, disease monitoring, and therapeutic guidance. EV-miRNAs offer stable and accessible indicators of patients’ inflammatory and metabolic status. For instance, elevated EV-miR-142-5p and miR-146a-5p might indicate an obesity-related inflammatory phenotype in MS, potentially guiding aggressive early intervention or lifestyle modifications. Additionally, these findings suggest therapeutic opportunities targeting specific miRNAs using miRNA mimics or inhibitors. MiRNA-based therapies, potentially delivered through engineered EVs or nanoparticles, represent innovative approaches to modulate neuroinflammation and metabolic dysregulation in MS. Preclinical studies in EAE models have demonstrated that manipulating miR-155 or enhancing miR-219 can ameliorate disease severity, supporting the feasibility of these strategies [85].

In conclusion, this study highlights the pivotal roles of EV-miRNAs in POMS, particularly emphasizing the amplifying effect of obesity on immune-metabolic disturbances through altered lipid metabolism and inflammation. These results establish a foundation for future validation of EV-miRNAs as biomarkers and for exploring miRNA-targeted therapeutics. Future research involving larger POMS cohorts and experimental models will be essential for confirming these mechanisms and translating them into personalized clinical strategies. Ultimately, integrating metabolic profiling with molecular characterization holds promise for improved MS outcomes, underscoring the importance of early intervention in managing obesity-related neuroinflammation.

### Limitations

Despite the robust methodological approach, our study has several limitations. Firstly, our sample size was small, limiting the statistical power to detect subtle differences and potentially reducing the generalizability of the findings. Larger cohorts are required to validate the observed EV-miRNA signatures and functional pathways. Secondly, this was a cross-sectional analysis; therefore, we could not assess longitudinal changes in EV-miRNA expression or determine causality between obesity and MS progression. Future longitudinal studies will be essential to explore the temporal dynamics of EV-miRNAs with disease evolution.

Additionally, although we carefully matched our POMS and control cohorts by age, sex, and BMI category, we did not account for potential confounders such as pubertal status, diet, physical activity levels, or medication effects, all of which may independently affect miRNA expression.

However, this information is being collected on our larger cohort for future studies. Furthermore, our study utilized circulating plasma EVs; thus, it remains unclear whether the identified EV-miRNAs precisely reflect CNS-specific processes or peripheral systemic responses. Integrating analyses of cerebrospinal fluid-derived EVs could provide a more direct assessment of CNS pathology.

Moreover, our bioinformatics analyses relied on computational prediction databases for miRNA target identification and pathway enrichment, which, despite extensive validation in the literature, might contain inaccuracies. Experimental validation of key predicted miRNA-mRNA interactions is necessary to establish their biological relevance conclusively.

Finally, while our data suggest that EV-miRNAs are promising biomarkers and therapeutic targets, translating these findings into clinical practice requires additional preclinical validation and mechanistic investigations in animal models or cellular systems to confirm their functional roles and therapeutic potential.

## Methods

### Ethics Approval

This study involves human participants and was approved by the University of Virginia (UVA) Institutional Review Board for Health Sciences Research (study ref ID: 20877, date: 08/14/2018). All participants provided informed consent prior to the start of any study-related procedures.

### Study participants

60 youth with POMS and 60 age-, sex-, and BMI-category matched controls were enrolled into a cross-sectional, prospectively recruited study at the University of Virginia. From this larger cohort, a small subset of 11 subjects (controls, n=6; POMS, n=5) was selected for inclusion in the current pilot study. Group BMI stratification followed CDC percentiles for healthy weight (BMIs ≤ 85th%tile) and overweight/obese (BMIs >85th%tile).

### Thrombin treatment for plasma defibrination

Human plasma samples were treated with thrombin to remove fibrinogen before EV isolation. Thrombin treatment was performed using a standardized protocol to facilitate the comparison of EV yield from defibrinated and non-defibrinated plasma. Specifically, 250 µL of human plasma was treated with 2 µL of thrombin (550 U/mL stock concentration; Cat# 605157, EMD Millipore, Burlington, MA, USA). The mixture was gently inverted several times to ensure uniform mixing and incubated at room temperature for 5 minutes to facilitate fibrinogen conversion to fibrin. Following incubation, the thrombin-treated plasma was centrifuged at 10,000 × g for 5 minutes to pellet the fibrin clot. The supernatant, representing the defibrinated plasma, was carefully transferred to a new sterile tube without disturbing the fibrin clot pellet.

### Ev isolation via tangential flow filtration (TFF)

EVs were isolated from both defibrinated and non-defibrinated plasma samples using tangential flow filtration (TFF) to efficiently concentrate and purify EVs. Plasma samples were initially filtered through 0.2 µm polyethersulfone (PES) syringe filters (MilliporeSigma, Burlington, MA, USA) to remove cells and debris. The filtered plasma was concentrated to a volume of 5 mL and adjusted to a final volume of 7 mL with sterile phosphate-buffered saline (PBS, pH 7.4), accounting for 2 mL of dead volume and 5 mL of liquid within the TFF tubing, protecting EVs from desiccation and ensuring consistent diafiltration.

TFF was performed using a self-assembled system equipped with a 500 kDa molecular weight cutoff (MWCO) hollow fiber filter (Repligen, Waltham, MA, USA). A peristaltic pump (Cole-Parmer, Vernon Hills, IL, USA) circulated the plasma at a constant flow rate of 35 mL/min. During diafiltration, PBS was continuously added to the reservoir at the same rate as permeate removal to maintain constant volume and effectively remove low-molecular-weight proteins, free nucleic acids, and other soluble contaminants.

Following TFF, the EV-enriched fraction (retentate, approximately 2 mL) was further concentrated using 30 kDa MWCO centrifugal filter units (MilliporeSigma Amicon Ultra Centrifugal Filter Unit, Burlington, MA, USA; Fisher Scientific, Pittsburgh, PA, USA) by centrifugation at 4000 × g for 30 min at 4 °C. The final EV samples were resuspended in 100 µL of sterile PBS for subsequent characterization and analysis.

### Microfluidic Resistive Pulse Sensing (MRPS)

The concentration and size distribution of EVs were analyzed using MRPS on the Spectradyne nCS1 instrument (Spectradyne, Torrance, CA, USA). Measurements were conducted using C-400 polydimethylsiloxane cartridges, which enable particle detection in a size range of approximately 65–400 nm in diameter. Data processing and analysis were performed using the nCS1 Data Analyzer software (Spectradyne, Torrance, CA, USA).

### Transmission Electron Microscopy (TEM) of plasma-derived EVs

To visualize plasma-derived EVs, TEM was performed following a standardized sample preparation protocol. TEM grids (Formvar/Carbon 200 mesh copper grids, Cat # FCF200-Cu; Electron Microscopy Sciences, Hatfield, PA, USA) were subjected to 1-minute plasma treatment before sample deposition. On a parafilm surface, two 20 µL droplets of deionized (DI) water and two droplets of UranyLess EM contrast stain (Cat # 22409; Electron Microscopy Sciences, Hatfield, PA, USA) were prepared.

For sample application, 10 µL of plasma-derived EVs were carefully pipetted onto the plasma-treated grids using a drop-casting method and allowed to incubate for 1 minute. Excess liquid was removed by gently blotting with filter paper. The grids were then sequentially washed by briefly dipping into a DI water droplet, followed by blotting with filter paper. This washing step was repeated with a fresh DI water droplet. The same process was applied for contrast staining, where the grids were incubated for 22 seconds in the UranyLess staining solution before blotting. To ensure complete drying, the prepared TEM grids were stored overnight in a designated grid box. Imaging was conducted using a Tecnai TF-20 transmission electron microscope (FEI Company, Hillsboro, OR, USA) operating at 200 kV to capture high-resolution images of the EVs.

### Western blotting

EV samples were lysed in radioimmunoprecipitation assay (RIPA) buffer (Thermo Scientific, Waltham, MA, USA) supplemented with protease and phosphatase inhibitor cocktails (Thermo Scientific, Waltham, MA, USA) and incubated on ice for 15 minutes. Protein concentrations were quantified using the Micro BCA™ Protein Assay Kit (Thermo Scientific, Waltham, MA, USA). Equal protein amounts were mixed with Laemmli sample buffer containing 2-mercaptoethanol (Sigma-Aldrich, St. Louis, MO, USA), briefly heated, and separated via sodium dodecyl sulfate-polyacrylamide gel electrophoresis (SDS-PAGE) on 4–20% Mini-PROTEAN® TGX Stain-Free gels (Bio-Rad, Hercules, CA, USA). Proteins were transferred to polyvinylidene fluoride (PVDF) membranes (Bio-Rad, Hercules, CA, USA). Membranes were blocked and incubated overnight at 4°C with primary antibodies against CD63, CD9 (Abcam, Cambridge, MA, USA) and Calnexin (Novus Biologicals, Centennial, CO, USA), diluted in Tris-buffered saline containing 0.1%

Tween-20 (TBS-T). After thorough washing, membranes were incubated for 1 hour at room temperature with horseradish peroxidase (HRP)-conjugated secondary antibodies. Protein bands were visualized using Clarity Max™ Western ECL Substrate (Bio-Rad, Hercules, CA, USA), and images were captured using the Bio-Rad ChemiDoc™ MP Imaging System.

### RNA extraction, miRNA sequencing and bioinformatics

RNA isolation, miRNA sequencing and bioinformatics were conducted by Creative Biostructures (Shirley, NY, USA).

Total RNA was extracted from the samples using the Trizol reagent (Invitrogen, CA, USA) following the manufacturer’s protocol. The quality and integrity of the extracted RNA were assessed using the Bioanalyzer 2100 (Agilent, CA, USA), ensuring a RNA integrity number (RIN) >7.0. The concentration of RNA was determined using a NanoDrop spectrophotometer (Thermo Fisher Scientific, MA, USA).

For small RNA library preparation, 1 µg of total RNA was used following the TruSeq Small RNA Sample Prep Kit protocol (Illumina, San Diego, USA). The adapter-ligated RNA was reverse transcribed to generate cDNA, which was then amplified using PCR. The libraries were purified and size-selected using polyacrylamide gel electrophoresis (PAGE) to enrich small RNA fragments. The purified libraries were quantified using the Qubit Fluorometer (Thermo Fisher Scientific, MA, USA) and assessed for quality using the Bioanalyzer 2100. The sequencing of the prepared small RNA libraries was performed using single-end 50 bp (SE50) sequencing on an Illumina HiSeq 2500 platform, following the standard protocol recommended by the manufacturer.

### Bioinformatics analysis

The raw sequencing reads were processed using an in-house bioinformatics pipeline (Creative Biostructures, USA). The initial steps included removal of adapter sequences, low-quality reads, and junk sequences, filtering out common RNA species including ribosomal RNA (rRNA), transfer RNA (tRNA), small nuclear RNA (snRNA), and small nucleolar RNA (snoRNA), and elimination of low-complexity sequences and repetitive elements. Subsequently, unique sequences of 18–26 nucleotides were aligned to [insert human genome version that was used] in miRBase 22.0 using BLAST to identify known and novel miRNAs. The alignment allowed for length variations at both the 3’ and 5’ ends and permitted a maximum of one mismatch within the sequence.

Sequences mapping to the mature miRNAs in hairpin arms were classified as known miRNAs. Sequences mapping to the opposite arm of a known miRNA precursor but not annotated as mature miRNA were classified as novel 3p- or 5p-derived miRNAs. To identify potential homologous miRNAs, unmapped sequences were aligned to other precursor miRNAs in miRBase 22.0. Those pre-miRNAs were further mapped to the reference genome using BLAST to determine their genomic locations. Unmapped sequences were then aligned to the reference genome, and potential miRNA hairpin structures were predicted using RNAfold software (http://rna.tbi.univie.ac.at/cgi-bin/RNAWebSuite/RNAfold.cgi).

The criteria for secondary structure prediction included the number of nucleotides in one bulge in the stem being ≤ 12, the number of base pairs in the stem region of the predicted hairpin being ≥ 16, a cutoff of free energy (kCal/mol ≤ -15), the length of the hairpin (up and down stems + terminal loop) being ≥ 50, and the length of the hairpin loop being ≤ 20. Additional criteria considered were the number of nucleotides in one bulge in the mature region being ≤ 8, the number of biased errors in one bulge in the mature region being ≤ 4, the number of biased bulges in the mature region being ≤ 2, the number of errors in the mature region being ≤ 7, and the number of base pairs in the mature region of the predicted hairpin being ≥ 12. Finally, the percentage of the mature sequence located within the stem region had to be at least 80%.

### Differential expression analysis

Differential expression analysis of miRNAs was conducted to identify significantly upregulated and downregulated miRNAs between controls versus POMS. Differences were compared using t-test with BH adjustment for multiple comparisons. Given our small sample size, we chose a less stringent p-value ≤ 0.1 to define statistically significant miRNAs, but also evaluated differences with p-values of 0.001, 0.01 and 0.05. FDR-adjusted q-values were also obtained.

### Volcano plot analysis

A volcano plot was generated to visualize the relationship between statistical significance and fold change of miRNA expression. Log2 fold change (log2FC) was plotted on the x-axis, while the negative logarithm of the *p-value* (-log10 p-value) was plotted on the y-axis. Significantly differentially expressed miRNAs were identified based on predefined thresholds for fold change and statistical significance, |log2FC| ≥ 1.2 and p-value ≤ 0.05. Red and blue points were used to indicate significantly upregulated and downregulated miRNAs, respectively, while non-significant miRNAs were shown in gray.

### Scatter plot analysis

To evaluate the correlation and overall distribution of miRNA expression levels between different sample groups, a scatter plot was generated. Normalized expression values from controls and POMS were plotted against each other to assess the global expression pattern. Each dot represented an individual miRNA, and a trendline was included to indicate the correlation between the datasets.

### Venn diagram analysis

To identify overlapping and unique EV-miRNAs between controls and POMS, a Venn diagram was constructed. The diagram was generated by comparing the differentially expressed miRNAs from both groups, and the number of common and unique miRNAs was displayed in their respective sections.

### Pathway analysis

To explore the biological functions and regulatory roles of the differentially expressed miRNAs, pathway enrichment analysis was performed using Kyoto Encyclopedia of Genes and Genomes (KEGG) and Gene Ontology (GO) databases. Target genes of differentially expressed miRNAs were predicted using publicly available databases such as TargetScan, miRDB, or miRanda.

The enriched pathways were identified based on statistical significance (p < 0.1), and the results were visualized using bar charts. Key biological processes, molecular functions, and cellular components associated with the identified miRNAs were analyzed to provide mechanistic insights into their roles in disease pathogenesis and physiological regulation.

## Data availability

All data generated or analyzed during this study are included in this manuscript and its supplementary information file. Further inquiries can be directed to the corresponding author.

## Author contributions

Conceptualization: MDH, ER, JNB, SMM

Study design and funding acquisition: ER, JNB, SMM

Investigation and conducted research: MDH, JNB

Organization and data analysis: MDH, SMM

Completed the original draft of the manuscript: MDH, SMM

Data interpretation, revision, and manuscript correction: MDH, ER, JNB, SMM

All authors approved the final version of the manuscript

## Additional information

Correspondence and requests for materials should be addressed to Senior Corresponding author.

## Data Availability

All data produced in the present study are available upon reasonable request to the authors.

## Acknowledgements

SMM was supported by intramural funding from the Abigail Wexner Research Institute at Nationwide Children’s Hospital and NINDS (5K12NS098482-05). ER was supported by National Institutes of Health (NIH) grants UG3/UH3TR002884 and U18TR003807. JNB was supported by NINDS (K23 NS116225). The content is solely the responsibility of the authors and does not necessarily represent the official views of the National Institutes of Health.

