## Supplementary information for "Extracellular vesicle miRNA signatures in pediatric onset-multiple sclerosis and obesity-driven immune and metabolic dysregulation"

700 Children’s Drive

Columbus, OH 43205, USA.

***Co-corresponding Author:** J. Nicholas Brenton, MD

Address:	University of Virginia

Division of Neurology, Division of Pediatric Neurology,

PO Box 800394

Charlottesville, VA 22908, USA.

**Supplementary Table 1: Differentially expressed EV-miRNAs Controls vs. POMS (p-value <0.1.)**

| Downregulated DEMs | FC | log2(FC) | p-value* |
| --- | --- | --- | --- |
| miR-99b-3p_R+1 | 0.68 | -0.55 | 5.62E-02 |
| miR-744-5p | 0.64 | -0.65 | 5.79E-02 |
| miR-543 | 0.43 | -1.22 | 6.48E-02 |
| miR-301a-3p | 0.76 | -0.4 | 6.58E-02 |
| let-7e-3p | 0.61 | -0.72 | 7.01E-02 |
| Upregulated DEMs |  |  |  |
| miR-6511a-3p_R+1 | 2.35 | 1.23 | 5.04E-02 |
| miR-548au-5p_R-1 | 1.68 | 0.75 | 5.13E-02 |
| miR-548am-5p_R-2 | 1.68 | 0.75 | 5.13E-02 |
| miR-133b_R-1 | 4.12 | 2.04 | 5.58E-02 |
| miR-6813-5p | 2.87 | 1.52 | 6.15E-02 |
| miR-548am-3p_R-1 | 7.99 | 3 | 6.17E-02 |
| miR-185-3p_R-1 | 1.43 | 0.52 | 6.46E-02 |
| miR-145-5p | 1.48 | 0.57 | 6.88E-02 |
| miR-548x-5p_L-2R+1 | 4.87 | 2.29 | 7.74E-02 |
| miR-548bc_R+1 | 2.39 | 1.26 | 7.90E-02 |
| let-7g-3p_R+1 | 3.35 | 1.74 | 7.94E-02 |
| miR-625-3p | 1.57 | 0.65 | 8.29E-02 |
| miR-548av-3p_L+2 | 1.37 | 0.45 | 8.87E-02 |
| miR-1273h-5p_R+2 | 1.27 | 0.35 | 9.04E-02 |

DEM, Differentially expressed miRNAs; FC, fold change; POMS, pediatric-onset multiple sclerosis

**Supplementary Table 2: Differentially expressed EV-miRNAs HC-NI vs. MS-NI**

**(p-value <0.1.)**

| Downregulated DEMs | FC | log2(FC) | p-value* |
| --- | --- | --- | --- |
| miR-514a-3p | 0.23 | -2.14 | 6.64E-02 |
| miR-1301-3p | 0.59 | -0.77 | 6.72E-02 |
| miR-223-5p_R+2 | 0.68 | -0.56 | 7.13E-02 |
| mir-1302-1-p5_1ss9AG | 0.24 | -2.08 | 7.36E-02 |
| miR-6837-3p | 0.11 | -3.15 | 7.72E-02 |
| miR-3120-3p | 0.66 | -0.6 | 7.77E-02 |
| miR-760_R+2 | 0.54 | -0.89 | 7.95E-02 |
| miR-3064-5p | 0.64 | -0.64 | 8.50E-02 |
| miR-139-3p | 0.57 | -0.81 | 9.04E-02 |
| miR-30d-3p | 0.62 | -0.69 | 9.10E-02 |
| miR-181b-5p_R-2 | 0.83 | -0.27 | 9.35E-02 |
| miR-148b-5p_L+1 | 0.81 | -0.3 | 9.80E-02 |
| miR-26b-3p_R+2 | 0.5 | -1.01 | 9.89E-02 |
| Upregulated DEMs |  |  |  |
| miR-1306-3p_R+4 | 1.67 | 0.74 | 5.92E-02 |
| miR-548av-3p_L+2 | 1.78 | 0.83 | 7.89E-02 |
| miR-4508_L+2R-1 | 3.44 | 1.78 | 8.53E-02 |
| miR-143-5p_R-1 | 2.22 | 1.15 | 8.88E-02 |
| miR-548bc_R+1 | 2.53 | 1.34 | 9.55E-02 |
| miR-5584-5p | 4.63 | 2.21 | 9.84E-02 |

DEM, Differentially expressed miRNAs; FC, fold change; HC-NI, healthy-weight pediatric control; MS-NI, healthy-weight pediatric-onset multiple sclerosis

**Supplementary Table 3: Differentially expressed EV-miRNAs PC-Ob vs. POMS-Ob (p-value <0.1.)**

| Downregulated DEMs | FC | log2(FC) | p-value* |
| --- | --- | --- | --- |
| miR-766-3p | 0.42 | -1.27 | 5.15E-02 |
| miR-1255b-5p | 0.32 | -1.64 | 5.45E-02 |
| miR-378a-3p | 0.61 | -0.71 | 5.66E-02 |
| miR-148a-5p | 0.62 | -0.68 | 6.44E-02 |
| miR-543 | 0.41 | -1.3 | 6.84E-02 |
| miR-151a-5p | 0.74 | -0.44 | 7.03E-02 |
| miR-1249-3p | 0.19 | -2.39 | 7.29E-02 |
| miR-5010-3p_R+1 | 0.6 | -0.75 | 7.30E-02 |
| miR-598-3p | 0.77 | -0.38 | 7.31E-02 |
| miR-30b-3p | 0.51 | -0.98 | 7.37E-02 |
| miR-17-3p | 0.61 | -0.72 | 7.40E-02 |
| let-7f-5p | 0.76 | -0.39 | 7.42E-02 |
| miR-5010-5p | 0.66 | -0.59 | 7.61E-02 |
| miR-328-3p | 0.26 | -1.95 | 8.05E-02 |
| miR-532-5p | 0.68 | -0.55 | 8.57E-02 |
| miR-301b-3p | 0.58 | -0.78 | 9.70E-02 |
| Upregulated DEMs |  |  |  |
| miR-874-3p | 2.62 | 1.39 | 5.12E-02 |
| miR-590-5p | 3.27 | 1.71 | 5.29E-02 |
| miR-361-3p | 2.87 | 1.52 | 6.87E-02 |
| miR-363-3p_R-1 | 1.42 | 0.51 | 7.28E-02 |
| miR-548d-3p | 2.81 | 1.49 | 7.64E-02 |
| miR-338-3p_R+1 | 1.57 | 0.65 | 7.96E-02 |
| miR-148b-5p_L+1 | 2.09 | 1.07 | 8.17E-02 |
| miR-193a-5p | 1.78 | 0.83 | 8.23E-02 |
| miR-126-5p | 1.96 | 0.97 | 8.77E-02 |
| mir-1302-1-p5_1ss9AG | 7.47 | 2.9 | 8.90E-02 |
| miR-7976_R+3 | 2.56 | 1.36 | 9.65E-02 |
| miR-223-5p_R+2 | 1.64 | 0.72 | 9.98E-02 |

DEM, Differentially expressed miRNAs; FC, fold change; PC-Ob, obese control; POMS-Ob, obese pediatric-onset multiple sclerosis

**Supplementary Table 4: Differentially expressed EV-miRNAs MS-NI vs. POMS-Ob (p-value <0.1.)**

| Downregulated DEMs | FC | log2(FC) | p-value* |
| --- | --- | --- | --- |
| miR-378a-3p | 0.58 | -0.79 | 5.08E-02 |
| miR-548h-3p_R-1_1ss4AG | 0.11 | -3.19 | 5.43E-02 |
| miR-98-5p | 0.75 | -0.42 | 5.93E-02 |
| miR-125b-5p | 0.71 | -0.5 | 6.10E-02 |
| miR-605-3p | 0.47 | -1.09 | 6.47E-02 |
| miR-4676-3p_L-1 | 0.41 | -1.28 | 6.82E-02 |
| miR-378d_R-1 | 0.46 | -1.11 | 6.95E-02 |
| miR-532-3p | 0.79 | -0.34 | 6.96E-02 |
| miR-4772-3p | 0.35 | -1.53 | 8.50E-02 |
| miR-4742-5p | 0.67 | -0.58 | 8.97E-02 |
| mir-1304-p5 | 0.52 | -0.94 | 9.26E-02 |
| miR-26b-5p_R+1 | 0.76 | -0.4 | 9.45E-02 |
| Upregulated DEMs |  |  |  |
| miR-221-3p | 1.71 | 0.77 | 5.72E-02 |
| miR-4732-5p | 2.31 | 1.21 | 6.40E-02 |
| miR-3615_R+2 | 1.94 | 0.96 | 7.29E-02 |
| miR-1307-3p_R+1 | 1.19 | 0.26 | 7.46E-02 |
| miR-4433a-3p | 4.45 | 2.15 | 7.61E-02 |
| miR-130a-3p | 1.71 | 0.78 | 9.48E-02 |
| miR-1301-3p | 1.67 | 0.74 | 9.51E-02 |
| miR-548u_R-1 | 3.93 | 1.98 | 9.59E-02 |
| miR-548aq-3p | 3.84 | 1.94 | 9.65E-02 |

DEM, Differentially expressed miRNAs; FC, fold change; MS-NI, healthy-weight pediatric-onset multiple sclerosis; POMS-Ob, obese pediatric-onset multiple sclerosis
